# Evaluating the impact of Hazelwood mine fire event on students’ educational development with Bayesian interrupted time series hierarchical meta-regression

**DOI:** 10.1101/2021.03.28.21254516

**Authors:** Caroline X. Gao, Jonathan C. Broder, Sam Brilleman, Tim C. H. Campbell, Emily Berger, Jillian Ikin, Catherine L. Smith, Rory Wolfe, Fay Johnston, Yuming Guo, Matthew Carroll

## Abstract

**Background:** Climate disasters such as wildfires, floods and droughts can introduce significant interruptions and trauma to impacted communities. Children and young people can be disproportionately affected with additional educational disruptions. However, evaluating the impact of disasters is challenging due to difficulties in establishing studies and recruitment post-disasters.

**Objectives:** With the increasing threat of climate change, we aimed to (1) establish a new analytical framework to evaluate the impact of climate disasters on academic achievement and (2) evaluate the impact of the 2014 Hazelwood mine fire (a six-week fire event in Australia).

**Methods:** Bayesian hierarchical meta-regression was developed to evaluate the impact of the mine fire using only aggregated school-level data from the standardised National Assessment Program-Literacy and Numeracy (NAPLAN) test. NAPLAN results and school characteristics (2008-2018) from 69 primary/secondary schools with different levels of mine fire-related smoke exposure were used to estimate the impact of the event. Using an interrupted time series design, the model estimated immediate effects and post-interruption trend differences with full Bayesian statistical inference.

**Results:** Major academic interruptions across NAPLAN domains were evident in high exposure schools in the year post-mine fire (greatest interruption in Writing: 11.09 [95%CI: 3.16-18.93], lowest interruption in Reading: 8.34 [95%CI: 1.07-15.51]). The interruption was comparable to a four to five-month delay in educational attainment and had not fully recovered after several years.

**Discussion:** Considerable academic delays were found as a result of a mine fire, highlighting the need to provide educational and community-based supports in response to future events. Importantly, this work provides a statistical method using readily available aggregated data to assess the educational impacts in response to other climate disasters.

**Graphical abstract:** 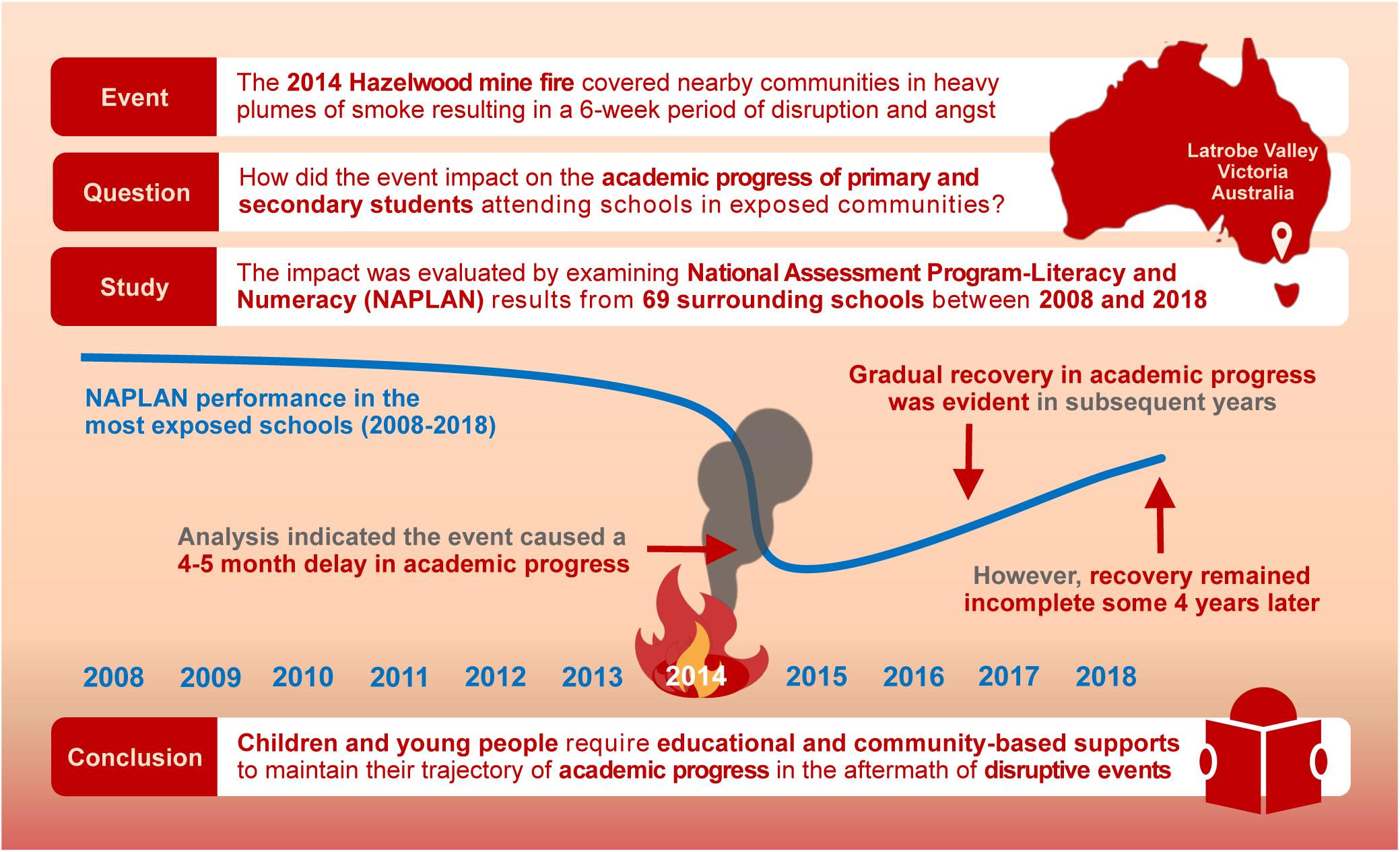

## 1 Introduction

Each year, climate disasters, such as wildfires, floods, droughts, affect 100 to 300 million people worldwide (Guha-Sapir et al., 2016). The frequency and severity of disasters such as wildfires have increased dramatically in recent years within the context of climate change (CRED, 2019; UNDRR, 2020; Xu et al., 2020). Children and adolescents are particularly vulnerable to experiencing difficulties in the aftermath of climate disasters due to physiological and developmental factors, as well as their dependence on others for care, protection and decision-making, associated with their age (Hoffman, 2008; Norris et al., 2002). The health, social and economic disruption of disasters impose substantial impacts on children and adolescents through factors such as trauma, illness, prolonged school interruption and reduced social support (Peek et al., 2018; Saylor, 1993).

It is well established that exposure to trauma in childhood is a potential pathway to later development of a range of psychological problems, such as anxiety, depression, fear, anhedonia, attention issues, aggressive behaviour and social withdrawal (Lubit et al., 2003). Indeed, posttraumatic stress disorder (PTSD), depression, anxiety and behavioural problems are widely reported in children and adolescents impacted by climate disasters (Kar, 2009; Kar and Bastia, 2006; McLaughlin et al., 2009; Terasaka et al., 2015). Wildfires have also been found to cause traumatic distress for children and adolescents (McFarlane et al., 1987, 1997; Yelland et al., 2010).

Exposure to disaster has the potential to result in long-term neurodevelopmental consequences for children and adolescents. Childhood trauma exposure is known to causing ongoing sensitivity to stress (De Bellis and Zisk, 2014), impair sequential development of brain structures and functions (De Bellis and Zisk, 2014; Perry, 2006) and cause difficulties in attention, memory and executive function (Buckley et al., 2000; Samuelson et al., 2010). These changes can be carried into adulthood and, in turn, increase risks of psychosocial problems throughout the lifespan (Caffo et al., 2005; De Bellis and Zisk, 2014; Edwardset al., 2003; Huh et al., 2014).

The psychological and neurodevelopmental impacts from trauma are important determinants of academic attainment in childhood and adolescence. The academic performance of students who have experienced trauma is generally poorer, and they are recognised to be at greater risk of early school dropout (Perfect et al., 2016; Romano et al., 2015). Although the predominant focus of childhood trauma research has been on the impacts of maltreatment, adverse academic outcomes have been similarly observed in children and adolescents following exposure to climate disaster (Gibbs et al., 2019; Perez-Pereira et al., 2012; Siriwardhana et al., 2013).

Climate disasters also impact children and adolescents through ongoing physical, educational, social and economical disruption. Schools may be forced to close or relocate for periods of time during and after disasters, causing interruptions and sometimes discontinuation in students’ academic programs, recreational activities, and social connections (Gibbs et al., 2019; Jaycox et al., 2007; Masten and Motti-Stefanidi, 2020; Peek and Richardson, 2010). Other physical, economic and social impacts, such as damage to property, loss of job opportunities, reduced incomes, increased distress levels and illness reported impacting adults (Xu et al., 2020; Broder et al., 2020; Norris et al., 2002; North and Pfefferbaum, 2013) can be potentially magnified in children and adolescents. However, the psychological and academic needs of children and adolescents have often been overlooked after disasters (McFarlane et al., 1987; Silverman and La Greca, 2002).

Therefore, there is a need to understand the impact of climate disaster on children and young people particularly under the increasing threat of climate change. However, evaluating the impacts of climate disaster on children and adolescents at a population-level is challenging, due to high cost, low response rates in surveys and lack of pre-disaster measurements necessary for evaluating changes (Kousky, 2016; Grievink et al., 2006; Tang et al., 2017). Administrative data such as the national standardised academic testing results (with participant rates of about 95%) can be used to evaluate the educational impact of disasters (Gibbs et al., 2019). However, accessing individual students’ academic records requires parental consent and complex ethics procedures, which can be a lengthy process and particularly challenging following disaster events.

These factors can lead to a lack of timely understanding of the degree and extent of climate disasters’ impacts, as well as how resources should be distributed to facilitate students’ coping and recovery and prevent long-term deficits and inequity. Therefore, there is an urgent need for establishing advanced methods that can use readily available aggregated educational data to evaluate the impact of climate disasters on children and adolescents.

In February 2014, a bushfire ignited the Morwell coal mine adjacent to the Hazelwood Power Station in the Latrobe Valley, Victoria, Australia and burned for approximately six weeks. Whilst the flames themselves did not directly threaten homes or lives, heavy smoke concentrations throughout the six-week period resulted in increased mortality, physical ill-health and psychological distress in the local community (Broder et al., 2020; Dimitriadis et al., 2021; Gao et al., 2020; Holt et al., 2021.; Ikin et al., 2020; Johnson et al., 2019). The event also caused considerable school disruption, including temporary closures and relocations (Berger et al., 2018; Ikin et al., 2020).

The Hazelwood Health Study (HHS; www.hazelwoodhealthstudy.org.au), established to evaluate the health and wellbeing impact of the mine fire (Ikin et al., 2020; Melody et al., 2020), conducted a school survey to evaluate the psychological outcomes of the mine fire on students attending schools in affected communities. A subsequent evaluation of the National Assessment Program-Literacy and Numeracy (NAPLAN) results of survey participants suggested academic delays in highly smoke-exposed areas (Berger, Gao, et al., 2020). However, the results from the study with the low participation rate, as is frequently the case in post-disaster studies, are at the risk of bias (Pullins et al., 2005; Salloum and Overstreet, 2012) and requires further evaluation. Aggregated school-level NAPLAN data were subsequently obtained to further consolidate our findings.

In this study, we developed a Bayesian interrupted time series hierarchical meta-regression model to evaluate the impact of the Hazelwood mine fire on academic performance. Using this method, instead of individual-level data, only aggregated school-level data from standardised academic tests is required for evaluating spatial and temporal profiles of community-wide traumatic events. Here, we have presented detailed analysis procedures and results to inform the conduct of research concerning educational outcomes after climate disasters.

## 2 Methods

### 2.1 Study design

#### 2.1.1 Selection of schools and classification of exposure risks

The modelled particulate matter exposure (PM∼2.5) during the mine fire period suggested that the town Morwell, a statistical area level 2 area (SA2, comparable to township) (ABS, 2011) closest to the mine fire experienced the highest smoke concentrations, which at times greatly exceeded national safety standards (Luhar et al., 2020). The wider Latrobe Valley (statistical area level 3 [SA3] area comparable to a local government area which includes Morwell) suffered a moderate level of exposure (Luhar et al., 2020). Due to wind direction, the nearby SA3 area of Wellington, with a similar socioeconomic profile to Latrobe Valley, had little to no exposure to the smoke. Therefore, Wellington was chosen as the comparison area for this analysis.

As the students would be exposed air pollution both at their school and at home, the smoke exposure of at individual school location was not used in the analysis. Instead, 69 primary and secondary schools in the area were classified into three mine fire exposure groups: Morwell (high exposure), the remainder of the Latrobe Valley (moderate exposure) and Wellington (no/low exposure), see Figure 1.

**Figure 1:**
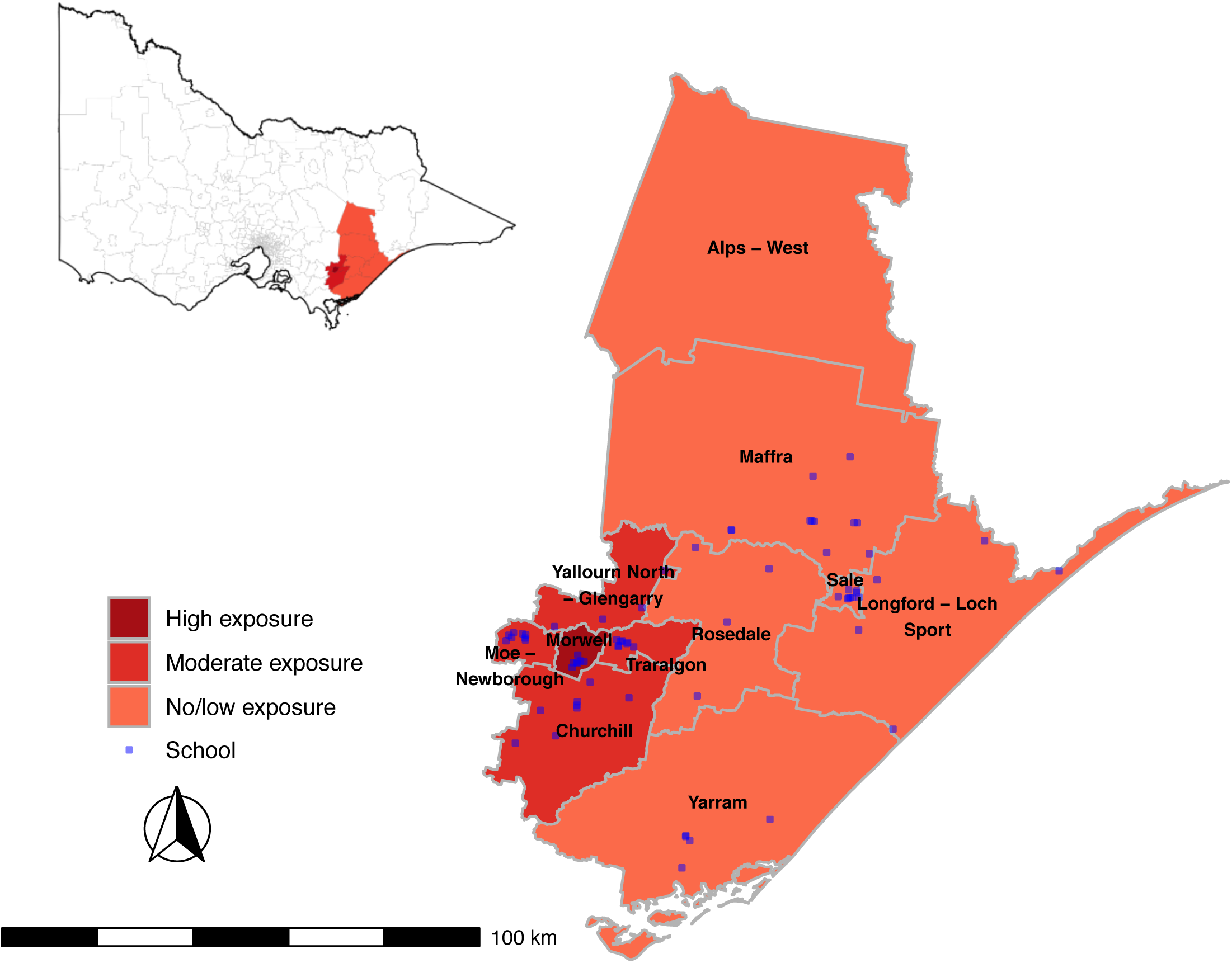
Map of mine fire exposed SA2 in Victoria. Superimposed dots are the location of schools in the impacted area

#### 2.1.2 NAPLAN and school profile data

In Australia, NAPLAN tests are conducted annually in May for Grade 3, 5, 7 and 9 students and NAPLAN examines educational domains of reading, writing, numeracy, spelling and grammar/punctuation (ACARA, 2019). As NAPLAN assesses incremental learning, students’ scores are expected to increase when they are re-tested at each two-year interval, from an average score of around 420 in Grade 3, to 580 in Grade 9 across individual domains (ACARA, 2019).

Aggregated school-level NAPLAN data (mean score and SE for each domain) were requested from the Australian, Curriculum, Assessment and Reporting Authority (ACARA) for all Victorian schools between 2008 and 2018. School profiles, including total enrolments, school sector (government, non-government), school type (primary, secondary), gender proportion and Index of Community Socio-Educational Advantage (ICSEA, a scale indicating socioeducational advantage of the school based on students’ family background information), were provided by ACARA for each year.

### 2.2 Statistical methods

School characteristics (in 2014) and NAPLAN participation rate (2008-2018) were first compared across the three exposure groups. School-level mean NAPLAN scores pre- and post-mine fire were visualised using box plots by exposure group for each educational domain and student grade level.

#### 2.2.1 Centring NAPLAN scores

NAPLNA tests were designed to represents growth of student’s achievement over time from Grade 3 to Grade 9 with scores increase for each test. As the test score can be interpreted in terms of academic progression, it was not standardized. However, to make results comparable across student’s grades and domains, all scores were first centred against the matching mean scores in regional Victoria (for the same year, grade and educational domain).

#### 2.2.2 Bayesian interrupted time series hierarchical meta-regression

Hierarchical two-level meta-regression models were carried out in a Bayesian modelling framework to estimate the association between mine fire exposure and centred NAPLAN scores. This approach directly model distributions of students’ performance within schools using mean scores and standard errors, which provides unbased estimates that are comparable with the model using individual-level records. Interrupted time series model using only means with linear regression instead of meta-regression will be inefficient (significant loss of statistical power) and also bias towards smaller schools (large schools with more students should be given more weights). Although hierarchical meta-regressions can be carried out in non-Bayesian framework, the complex model structure can introduce convergence difficulties and problematic confidence intervals (van Houwelingen et al., 2002). In the Bayesian framework prior information can be also incorporated.

The modelling framework is illustrated in Figure 2. The first level random effects (random intercepts) were modelled as student cohorts, e.g., the cohort of students progressing from Grade 3 in 2014 to Grade 5 in 2016 at the same school. The nested second level random effects (random intercepts) were modelled at the school-level. The centred mean NAPLAN score at Grade *g* level for cohort *c* students in given school *s* was modelled as follows:

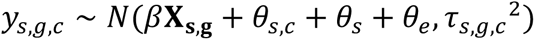

**Figure 2.**
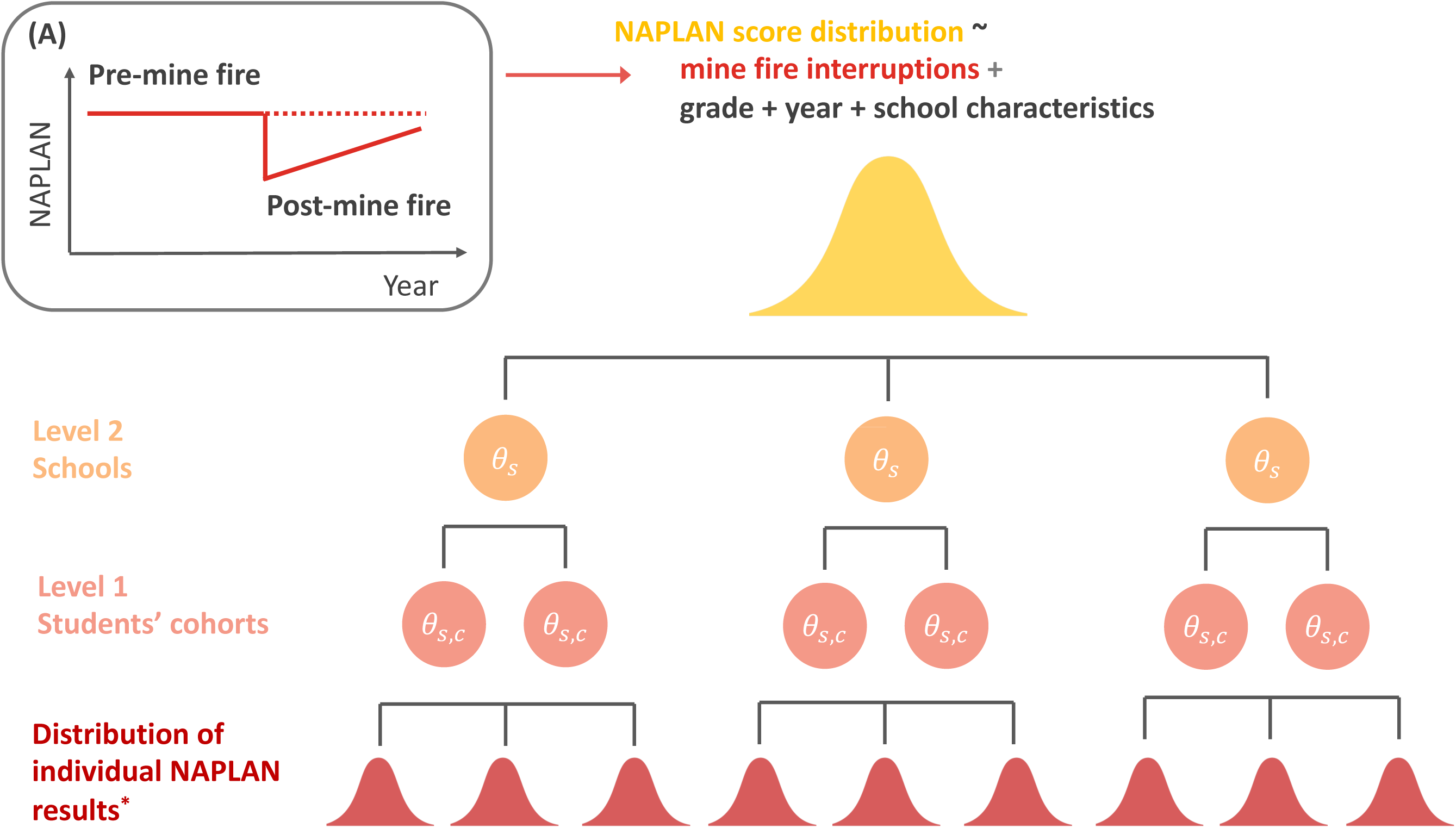
Illustration of Bayesian interrupted time series hierarchical meta-regression *Means and SE of test results each year for individual students’ grade level at each school. The subgraph (A) represents a theoretic model of the mine fire interruption effect

The terms *θ_s_* and *θ_s,c_* are the random effects for the school *s* and cohort *c* in that school, respectively. *θ_e_* is the random error term. We assume 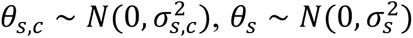 and *θ_e_* ∼ 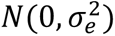. *τ_s,g,c_* is the standard error of the mean NAPLAN score of the given school, student cohort and grade level (input data). The matrix **X_s,g_** for fixed effects includes potential confounding factors: school characteristics (ICSEA, total enrolments, percentage of girls, school sector), grade level, long-term trend (year) as well as mine fire exposure effect variables detailed below.

The mine fire exposure effect was evaluated using the interrupted time series design (Bernal et al., 2017) with a time-specific interruption variable to capture the immediate effect of the mine fire on that year’s NAPLAN results, and a change in trend from pre- to post-mine fire (an interaction between the time interruption variable and year), see Figure 2(A). It was hypothesised that there would be no interruption effect in the no/low exposure group. Mine fire interruption effects and post interruption trend differences were evaluated separately for the high and moderate exposure groups, summarised as follows:

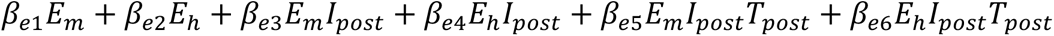

where *E_m_* and *E_h_* are indicator variables for moderate and high exposure. *I_post_* is the indicator variable for pre or post-mine fire (0 for 2008-2013 and 1 for 2014-2018). *T_post_* is the post-mine fire time variable (0 for 2008-2014, 1 for 2015, 2 for 2016 etc.). Therefore, *β_e_*_1_ and *β_e_*_2_ can be interpreted as the prior-mine fire differences when comparing moderate and high exposure schools with no/low exposure schools. These two coefficients are subsequently referred to as the fixed intercepts. The coefficients *β_e_*_3_ and *β_e_*_4_ can be interpreted as the mine fire interruptions effect (relative to the students’ developmental trajectories) for moderate and high exposure schools, respectively. The coefficients *β_e_*_5_ and *β_e_*_6_ are the post-mine fire trend differences compared with the trend before the mine fire. Other than *β_e_*_1_, *β_e_*_2_…*β_e_*_6_, the vector of coefficients, *β*, also contains coefficients of other confounding variables detailed above.

#### 2.2.3 Estimation and modelling framework

Estimation was conducted by Markov chain Monte Carlo (MCMC) sampling implemented in the Stan (Stan Development Team, 2020) programming language via RStan package (Stan Development Team, 2020). Stan is a platform for high-performance full Bayesian statistical inference using “No-U-Turn Sampler” (NUTS) (Hoffman and Gelman, 2014). Weakly informative prior distributions were used for standard deviations (SDs) of random effects, namely, *σ_s,c_* ∼ *TN*(10, 5^2^), *σ_s_* ∼ *TN*(10, 5^2^) and *σ_e_* ∼ *TN*(10, 5^2^), where TN is the truncated normal distribution with support over the range [0, *infinity*). The mean of 10 for prior distributions of the random effects was chosen based on a preliminary exploratory evaluation of all Victorian schools. Weakly informative prior distributions *N*(0, 50^2^) were used for all fixed effect parameters as non-informative priors base on uniform distributions can lead to a range of model fitting issues (Gelman et al., 2017; Stan Development Team, 2020).

Separate models were estimated for each testing domain. Results were reported as the posterior mean of estimated coefficients (4 Monte Carlo chains with 2000 iterations each), 95% credible interval (CI) and the probability of estimated coefficients (*β*) being greater or less than 0. Predicted centred NAPLAN score for schools were calculated and visualised for each testing domain and exposure group using line plots.

#### 2.2.4 Sensitivity analyses

A range of sensitivity analyses were undertaken to test the robustness of results, which included using different prior distributions and excluding cohort random effects. Also, two schools in Morwell were relocated during the mine fire event and remained at their relocation sites for an extended period afterwards; hence sensitivity analyses were conducted excluding the two relocated schools. Code for fitting the models using synthetic data is provided in the Supplementary Material I.

## 3 Results

### 3.1 School characteristics

The characteristics of schools across exposure groups are presented in Table 1. ICSEA socio-educational advantage scores were lower for schools in the high exposure group (Morwell) compared to schools in the moderate and no/low exposure groups. Other school characteristics, including the percentage of girls, number of students, school sector and NAPLAN test participation rates were comparable between exposure groups.

**Table 1:** School profile (in the year 2014) and NAPLAN participation rate by exposure area

|  | Overall<br>(N = 69) | No to low<br>exposure<br>(N = 34) | Moderate<br>exposure<br>(N = 28) | High<br>exposure<br>(N = 7) |
| --- | --- | --- | --- | --- |
| ICSEA | 963 (938, 998) | 974 (954, 998) | 962 (942, 1,006) | 922 (906, 944) |
| Proportion of girls | 0.49 (0.45, 0.51) | 0.49 (0.45, 0.51) | 0.49 (0.46, 0.51) | 0.47 (0.44, 0.51) |
| Total enrolments | 153 (79, 316) | 108 (47, 254) | 165 (118, 340) | 204 (142, 280) |
| School type |  |  |  |  |
| Combined | 1 (1.4%) | 1 (2.9%) | 0 (0%) | 0 (0%) |
| Primary | 60 (87%) | 29 (85%) | 25 (89%) | 6 (86%) |
| Secondary | 8 (12%) | 4 (12%) | 3 (11%) | 1 (14%) |
| School sector |  |  |  |  |
| Government | 53 (77%) | 26 (76%) | 22 (79%) | 5 (71%) |
| Non-government | 16 (23%) | 8 (24%) | 6 (21%) | 2 (29%) |
| Participation rate |  |  |  |  |
| 2008 | 0.97 (0.93, 1.00) | 0.97 (0.93, 1.00) | 0.97 (0.94, 1.00) | 0.96 (0.92, 0.99) |
| 2010 | 0.94 (0.88, 0.98) | 0.97 (0.89, 1.00) | 0.94 (0.91, 0.96) | 0.82 (0.80, 0.85) |
| 2012 | 0.94 (0.91, 1.00) | 0.97 (0.92, 1.00) | 0.93 (0.90, 0.96) | 0.92 (0.88, 0.95) |
| 2014 | 0.96 (0.90, 0.98) | 0.96 (0.95, 0.98) | 0.93 (0.88, 0.97) | 0.94 (0.92, 0.96) |
| 2016 | 0.94 (0.91, 0.97) | 0.95 (0.91, 0.97) | 0.94 (0.91, 0.97) | 0.92 (0.88, 0.93) |
| 2018 | 0.94 (0.89, 0.97) | 0.94 (0.88, 0.97) | 0.94 (0.90, 0.95) | 0.97 (0.91, 0.99) |

Note: Statistics presented are median (IQR) and n (%)

### 3.2 NAPLAN score pre-and post-mine fire

Distributions of NAPLAN scores pre- and post-mine fire for each domain and grade level were plotted by exposure group, which are presented in Figure 3. While there was a general trend across all exposure groups for NAPLAN performance to decline post-mine fire, this was greater in higher exposure schools.

**Figure 3:**
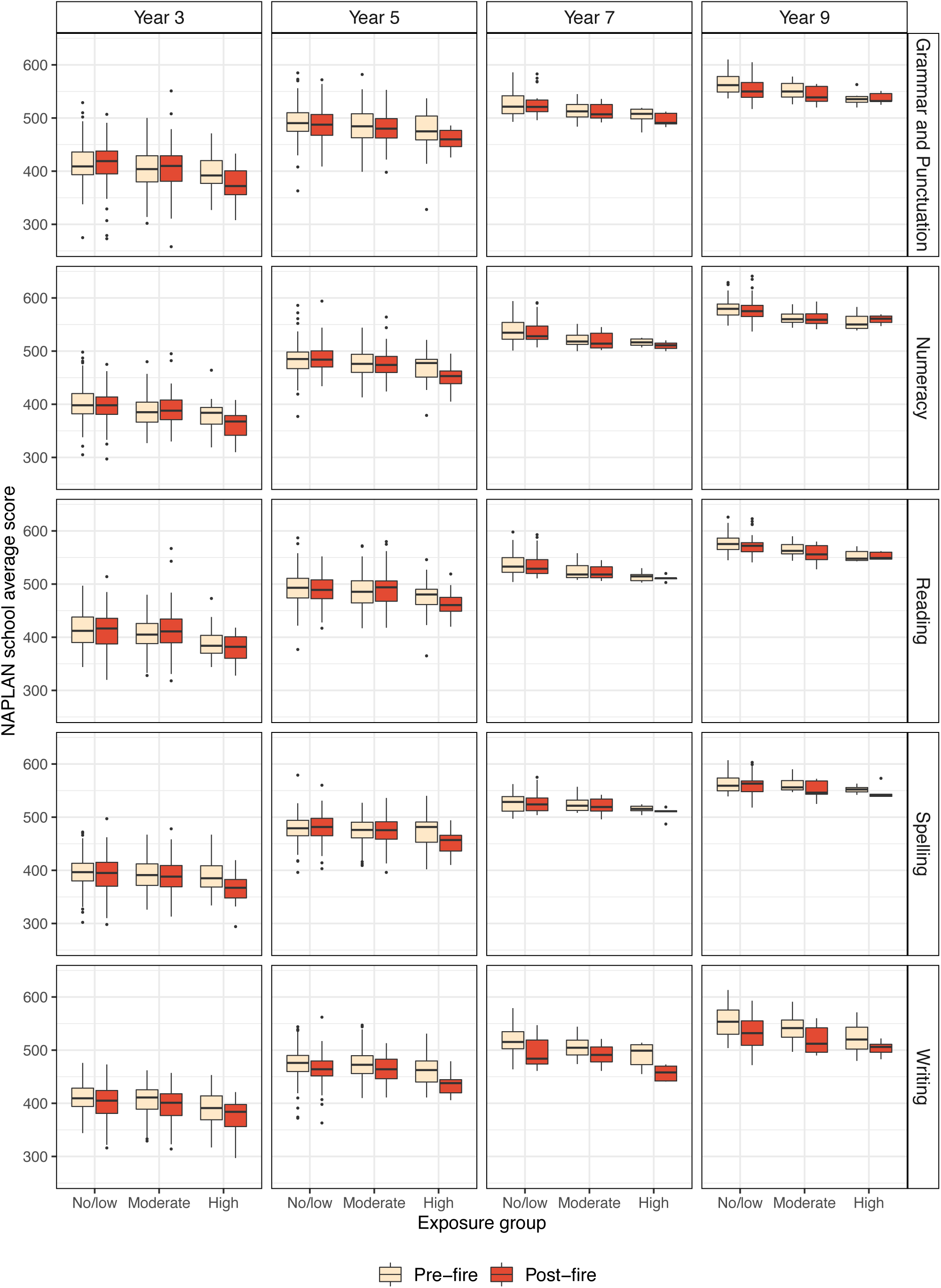
Box plot of school mean NAPLAN score for pre-mine fire period and post-mine fire period by exposure group for each domain and grade level

### 3.3 Bayesian hierarchical meta-regression models

Results from the Bayesian hierarchical meta-regression models are presented in Table 2 and Figure 4. Compared with the Victorian regional average, there was an estimated downward trend across all domains of testing for the schools in the three exposure groups (see Figure 4 and Table S1 to S5 in Supplementary Material II). As shown in Table 2, NAPLAN scores were found to be similar between schools in the moderate and no/low exposure group pre-mine fire after controlling for other confounding factors (e.g., there was no evidence that the fixed intercept mode coefficients for moderate exposure differed from 0). However, pre-mine fire NAPLAN scores were estimated to be lower in schools in the high exposure group for most domains when compared with no/low exposure schools (fixed intercept for high exposure ranged between -3 to -14, Table 2).

**Figure 4:**
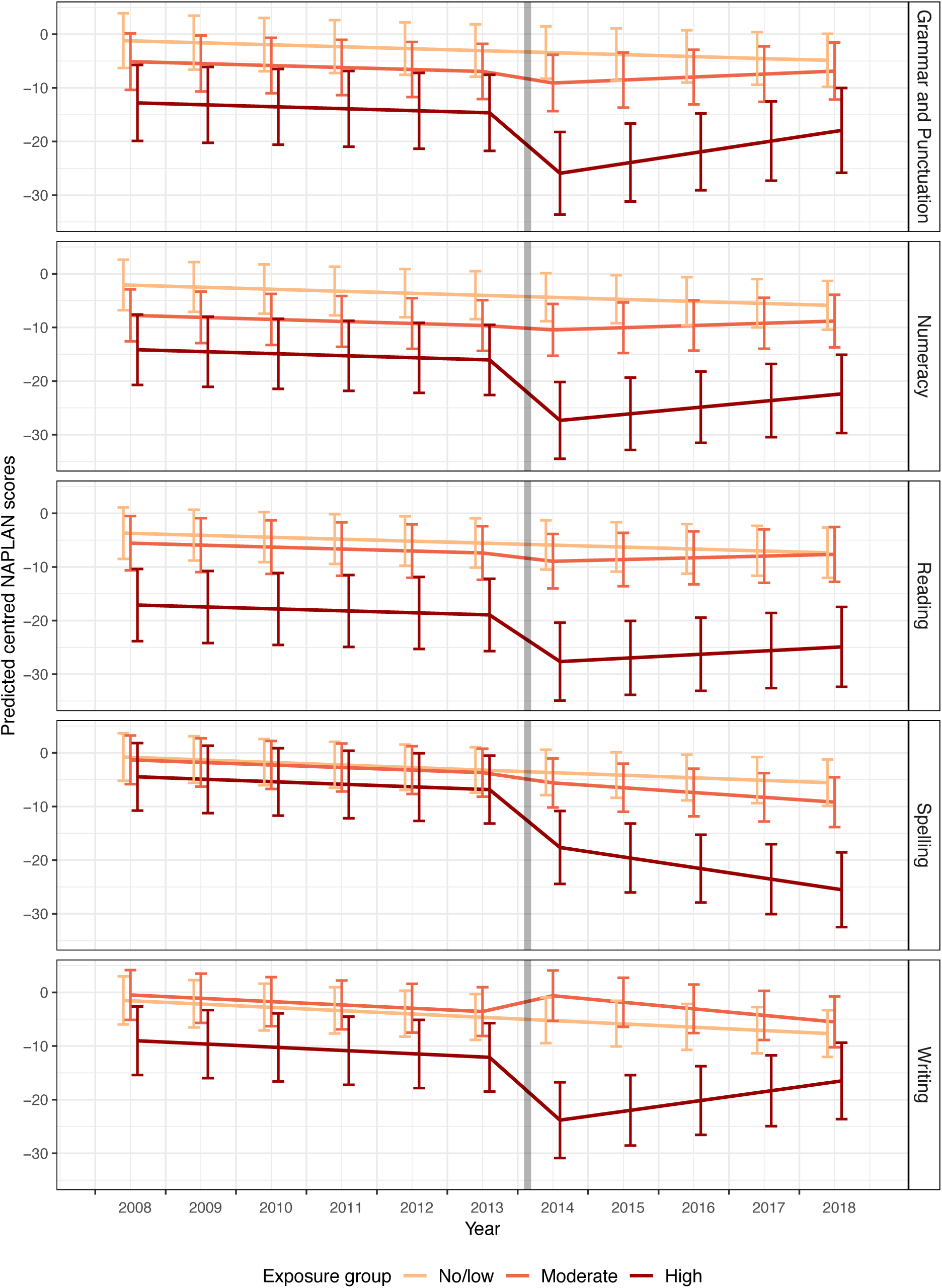
Trends of predicted centred NAPLAN scores (predicted margins when all covariates fixed at reference values) with error bars by exposure group for each academic domain. Note: centred NAPLAN scores represent score differences of schools compared with the regional Victorian average scores in the matching year, grade level and domain; the grey line indicates the time of the mine fire

**Table 2:** Estimated intercept, mine fire interruption effect and post-mine fire trend difference for moderate and high exposure schools estimated from Bayesian hierarchical meta-regression models

|  | Moderate exposure |  |  | High exposure |  |  |
| --- | --- | --- | --- | --- | --- | --- |
| | $\beta$ (95%CI) | $P(\beta < 0)$ | $P(\beta > 0)$ | $\beta$ (95%CI) | $P(\beta < 0)$ | $P(\beta > 0)$ |
| **Grammar and Punctuation** |  |  |  |  |  |  |
| Fixed intercept | -3.9 (-13.13, 5.31) |  | 0.20 | -11.61 (-25.28, 2.18) |  | 0.05 |
| Mine fire interruption effect | -1.78 (-5.73, 2.13) |  | 0.19 | -10.91 (-18.68, -3.08) |  | 0.004 |
| Post-mine fire trend difference | 0.92 (-0.51, 2.35) | 0.11 |  | 2.37 (-0.73, 5.37) | 0.06 |  |
| **Numeracy** |  |  |  |  |  |  |
| Fixed intercept | -5.66 (-13.94, 2.60) |  | 0.08 | -12.07 (-24.26, 0.23) |  | 0.028 |
| Mine fire interruption effect | -0.44 (-4.20, 3.28) |  | 0.41 | -10.9 (-17.98, -3.67) |  | <0.001 |
| Post-mine fire trend difference | 0.79 (-0.56, 2.13) | 0.13 |  | 1.61 (-1.21, 4.43) | 0.13 |  |
| **Reading** |  |  |  |  |  |  |
| Fixed intercept | -1.87 (-10.19, 6.39) |  | 0.33 | -13.41 (-26.26, -0.19) |  | 0.024 |
| Mine fire interruption effect | -1.18 (-4.84, 2.57) |  | 0.27 | -8.34 (-15.51, -1.07) |  | 0.013 |
| Post-mine fire trend difference | 0.69 (-0.61, 2.01) | 0.15 |  | 1.05 (-1.63, 3.85)) | 0.23 |  |
| **Spelling** |  |  |  |  |  |  |
| Fixed intercept | -0.52 (-8.45, 7.05) |  | 0.45 | -3.68 (-16.16, 8.34) |  | 0.27 |
| Mine fire interruption effect | -1.44 (-4.91, 2.09) |  | 0.21 | -10.31 (-17.39, -3.38) |  | 0.003 |
| Post-mine fire trend difference | -0.42 (-1.68, 0.85) | 0.74 |  | -1.49 (-4.20, 1.21) | 0.86 |  |
| **Writing** |  |  |  |  |  |  |
| Fixed intercept | 1.01 (-6.83, 8.71) |  | 0.61 | -7.53 (-19.63, 4.62) |  | 0.11 |
| Mine fire interruption effect | 3.56 (-0.52, 7.55) |  | 0.96 | -11.09 (-18.93, -3.16) |  | 0.003 |
| Post-mine fire trend difference | -0.6 (-2.01, 0.82) | 0.8 |  | 2.44 (-0.60, 5.50) | 0.06 |  |

Note: All regression coefficients ( $\beta$ ), 95% Credible Intervals (95% CI), and posterior probabilities of coefficients under or above 0 [ $P(\beta < 0)$ , $P(\beta > 0)$ ] were estimated from multivariate Bayesian hierarchical meta-regression models, controlling for school-level confounders including ICSEA, total enrolments, percentage of girls, school sector, grade level, long-term. Mine fire interruption effects and post-mine fire trend differences were estimated using the interrupted time series design.

Table 2 also shows that there were substantial interruption effects post-mine fire in high exposure schools across all academic domains, with estimated mean score reductions of: -10.91 (95%CI: - 18.68, -3.08) for grammar and punctuation; -10.9 (95%CI: -17.98, -3.67) for numeracy; -8.34 (95%CI: -15.51, -1.07) for reading; -10.31 (95%CI: -17.39, -3.38) for spelling and -11.09 (95%CI: -18.93, -3.16) for writing. Typically, NAPLAN scores increase about 27 points per year (ACARA, 2019), which means that the delay in educational attainment in high exposure schools was equivalent four to five months.

After the initial drop in academic performance subsequent to the mine fire, there was evidence that writing and grammar and punctuation scores began to recover (positive slope) in high exposure schools, see Figure 4. However, there was no such improvement post-mine fire for spelling in high exposure schools (estimated slope of -1.49 post mine fire; 95%CI: -4.20, 1.21). Also, the predicted centred NAPLAN scores in 2018 were found to be lower compared with the pre-mine fire period in all academic domains for high exposure schools (see Figure 4), which indicates incomplete recovery five years post-mine fire.

### 3.4 Sensitivity analysis

The choice of prior distributions for the SDs of random effects were found to have little impact on results; however, using a weakly informative prior distribution reduced the time taken to fit the Bayesian models compared with using a non-informative prior distribution. Sensitivity analysis excluding the cohort random effects (only the school level clustering is considered) produced very similar results except for a slightly larger mine fire interruption effect for high exposure schools (see Table S6). Models excluding the two relocated schools showed results that were consistent with all schools, but with slightly smaller interruption effects (see Table S7). This suggests that school relocation might have had an adverse impact on NAPLAN performance in addition to the mine fire exposure. It’s also possible that the adverse effects were stronger in the relocated schools which were closest to the mine fire.

## 4 Discussion

This study presents an innovative method to evaluate the impact of disasters on students’ academic performance using only readily accessible aggregated school-level data. Results suggest that the Hazelwood mine fire had a major impact on academic performance in schools located within areas highly exposed to the smoke during the event. The impact of the event on academic achievement was consistent across all NAPLAN testing domains in high exposure schools, which is comparable with four to five months delay in educational attainment. While there was some recovery in academic performance in high exposure schools across all academic domains except for spelling, performance levels remained below those seen prior to the mine fire some four years afterwards. Given academic underachievement can lead to unemployment, social and economic disadvantage, and ill-health later in life (Bowman et al., 2017), it is critical that impacts of climate disaster on the academic progression of students are recognised and remedied.

### 4.1 Links between mine-fire and academic performance

The impact of the mine fire on academic performance may be attributable to a combination of factors. The mine fire introduced disruption to day-to-day operations in nearby schools (Berger et al., 2018). Two schools were relocated causing direct educational interruptions as well as threats to student’s sense of safety and security (Berger et al., 2018). The mine fire also led to ongoing distress experienced by students, teachers and parents in the smoke affected community post the mine fire (Berger et al., 2018; E. Berger, Maybery, et al., 2020; Broder et al., 2020; Maybery et al., 2020). Hazelwood mine fire-related air pollution has also been linked to a range of adverse physical health effects such as respiratory morbidity (Gao et al., 2020; Holt et al., 2021; Johnson et al., 2019; Shao et al., 2020). More broadly, the community wellbeing was also negatively impacted with local residents reporting a loss of trust in authorities and feeling abandoned with little support provided (Yell et al., 2019a). The subsequent closure of the Morwell mine and Hazelwood power station some three years after the fire occurred introduced additional social and economical challenges in an already disadvantaged community (VCOSS, 2015; Yell et al., 2019b). These factors may have impacted students’ learning abilities both directly and indirectly.

Potential links between air pollution and cognitive development, educational and behavioural outcomes in children is a rapidly increasing area of research. Associations have been found between chronic ambient air pollution and cognitive function as well as educational attainment (Clifford et al., 2016; Forns et al., 2017; Marcotte, 2017). A recent study suggested a link between prenatal airborne polycyclic aromatic hydrocarbons (PAH) exposure, which was also found in Hazelwood mine fire smoke (Reisen et al., 2017), and academic underachievement, particularly in spelling, during early adolescent years (Margolis et al., 2021). However, we are not aware of previous research evaluating associations between medium-duration air pollution events and long-term academic outcomes. Although our study design cannot delineate the relative contributions of all possible factors such as psychological trauma, disruption to schooling, or air pollution exposure, it indicates a possible linkage that warrants further investigation.

### 4.2 Improvements post-mine fire

Gradual improvements in academic performance were observed across most domains post-mine fire among students from high exposure schools. This may be a result of a slow recovery both at the individual and community level. Increases in school funding from the Victorian Government to upgrade school facilities and a range of implemented government strategies focused on improving health, wellbeing and access to services in the impacted community, may have assisted the recovery process (DTF, 2015; IGEM, 2017). Student’s performance in the spelling domain continued to deteriorate post-mine fire, which may be related idiosyncrasies in English language orthography (Babayiğit and Stainthorp, 2011; Barry and Seymour, 1988), or difficulties in teaching spelling (Adoniou, 2014; Bosman and Van Orden, 1997; Treiman, 2018). As difficulties in spelling may persists and lead to arrested writing development subsequently (Graham and Santangelo, 2014), further investigation is needed.

### 4.3 Comparison with other studies

Although a theoretical link between disasters and educational outcomes has been well-established (Peek, 2008), most studies have been limited to evaluations of how disasters impacted school attendance or drop-out rates (Perez-Pereira et al., 2012; Pietro, 2018; Siriwardhana et al., 2013) and academic delays have rarely been investigated. One study suggested more than 75% of the African American children evacuated in response to Hurricane Katrina (which resulted in the loss of more than 1,300 lives, 800,000 homes and 110 schools) experienced a decline in grades (Peek and Richardson, 2010). To our knowledge, only Gibbs and colleagues (2019) have previously evaluated the impact of disasters using national standardised academic performance tests. Their study found that exposure to the 2009 Black Saturday bushfire in Australia (loss of 173 lives, 2,000 homes and 3 schools) was associated with delays in academic achievement from Grade 3 to 5 in reading and numeracy but not in writing, spelling, and grammar domains (Gibbs et al., 2019). However, only academic progressions of a single cohort of students (who were in Grade 3 in 2011 and were aged around 7 at the time of the event) were evaluated. Our study, on the other hand, has utilized a time series design to assess academic achievement across all grade-levels completing NAPLAN testing and incorporated more historical data, which sheds further light on academic impacts caused across age groups.

### 4.4 Strength and limitations

This paper also provides an easily adaptable method to evaluate the impact of different types of community-level traumatic exposures (e.g., disasters and disease outbreaks) on school-aged children’s and adolescents’ academic progression using only aggregated school-level data. The method that has been developed has a number of strengths. Firstly, it enables the evaluation of the spatial and temporal profile of the impact, which could be used to inform policy and resource allocation in academic settings after disasters.

Secondly, the method can be applied using only summary statistics at the school-level to obtain unbiased estimates without losing substantial statistical power to detect meaningful differences. Although only 69 schools were included in our analysis, the study is effectively powered with over 30,000 students who have studied in these schools from 2008 to 2018. Accessing aggregated data is a particularly useful approach to research in this area as it circumvents the challenges of attempting to recruit individuals in communities post-disaster who are already burdened and traumatized as a result of their experiences, and is also a less resource-intensive approach. Similar approach can also be used without the interrupted time series design to study the associations between student’s education performance and other environmental factors such as air pollution.

Lastly, the interrupted time series design also facilitates comparison between academic outcomes pre- and post-disaster to identify changes in trends that may be attributable to the event. This approach can be easily adopted to provide information for teachers, schools and education departments to plan and implement educational modifications and accommodate students’ additional needs post-disaster.

This study also has recognised limitations. The model implemented assumes that student NAPLAN scores within schools were normally distributed, which could be unrealistic if, for example, distributions were skewed or the number of students are small. Although the number of schools in the high exposure group was modest, most of these schools had over 150 students enrolled, so the mine fire interruption effects identified are likely to be robust. Random slopes for schools, an extension of the model applied, was not considered due to the unwarranted increase in modelling complexity with limited data. Random slope and intercept models can be used where there are more schools and time points available. Furthermore, the aggregated nature of the data prohibited evaluating the contribution of individual-level risk factors to the academic outcomes observed. More detailed region-specific data on other risk factors, such as service availability, were not available for this analysis but in principle could be included in the proposed model.

## 5 Conclusion

This study provides evidence indicating that an extended air pollution event resulted in a delay in academic performance across multiple educational domains, which had not fully recovered several years after the event occurred. While most research to date has focused on the educational impact of maltreatment, or of major disasters causing significant loss of property and life, the current study shows that a community-wide traumatic event posing a minimal immediate risk to life and property can also have considerable long-term educational impacts. This finding highlights the substantial vulnerability of children and adolescents and the need to respond to community-wide disaster events, providing targeted support during and following the event, in the hope of preventing or ameliorating any educational impacts.

This paper provides a novel statistical method for using readily available aggregated data to assess educational impacts of disasters. Implementing research programs post-disaster is enormously challenging. Accordingly, an approach that enables accurate and timely assessment of educational impacts without impost on the community is invaluable. This model could be applied to investigate the effect of other extended events on academic achievement, for instance, the COVID-19 pandemic, which has impacted students’ access to, and delivery of, schooling worldwide.

## Data Availability

The data underlying this article were provided by Australian, Curriculum, Assessment and Reporting Authority (ACARA). Data will be shared on request to the corresponding author with the permission of ACARA. However, a synthetically generated data set based on the source data for the tutorial part of the paper is available at https://github.com/CarolineXGao/NAPLAN_
impact.

## Ethics approval

All procedure of the study was approved by the Monash University Human Research Ethics (project number: 5834) and the Victorian Department of Education and Training.

## Acknowledgements

We wish to thank the Latrobe Valley and Gippsland communities for their support and participation in the Hazelwood Health Study. We also like to acknowledge Prof Rob Hyndman for his generous sharing of the Rmarkdown LaTex template (https://github.com/robjhyndman/MonashEBSTemplates) for writing this paper in the Rmarkdown environment.

## Data availability statement

The data underlying this article were provided by Australian, Curriculum, Assessment and Reporting Authority (ACARA). Data will be shared on request to the corresponding author with the permission of ACARA. A synthetically generated data set based on the source data for the tutorial part of the paper is available at https://github.com/CarolineXGao/NAPLAN_impact.

## Supplementary material I - Tutorial for Bayesian interrupted time series hierarchical meta-regression

Import library packages

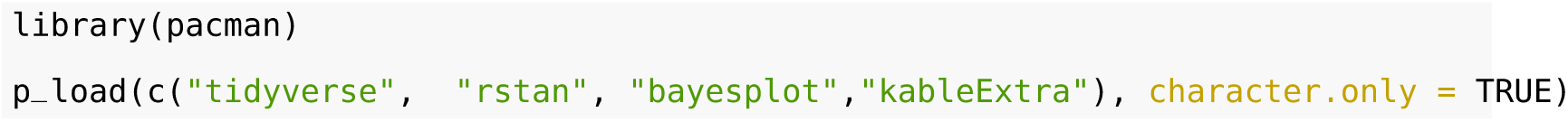

### Import data

The data as well as the analysis code used in this tutorial can be directly downloaded from Github repository: https://github.com/CarolineXGao/NAPLAN_impact.

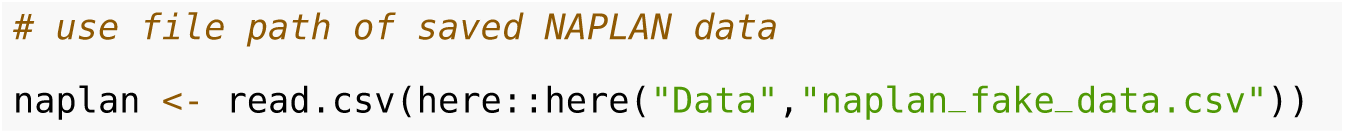

The variables in the data set are:

| Variable name | Details |
| --- | --- |
| ID | School ID |
| Cohort_ID | Cohort ID, which reflects the same cohort of students during NAPLAN assessment (e.g. grade 3 2003, grade 5 2005, grade 7 2007, grade 9 2009) |
| Year | Year of NAPLAN assessment |
| Grade | Grade of students who completed NAPLAN assessment |
| Domain | Domain of NAPLAN assessment, this domain is grammar and punctuation |
| ICSEA | Index of Community Socio-Educational Advantage per school and year |
| N_student | Number of students enrolled in the school in the year of NAPLAN test |
| P_girls | Proportion of female students in the school and year of NAPLAN test |
| Government | Whether the school is a government school |
| Exposure_group | Location of school as a proxy for coalmine fire exposure: Morwell schools - High exposure, non-Morwell Latrobe Valley schools - Moderate exposure, and Wellington Schools - No/low exposure |
| ScoreDif | Mean NAPLAN score of the school - Victorian regional schools mean score for the matching year and grade level (mean NAPLAN score difference) |
| SE | Standard error of mean NAPLAN score for the school, year and grade level |

### Modelling

#### Prepare data for modelling

A number of variables need to be changed for Bayesian modeling with Stan. Categorical variables, including grade (Grade) and exposure group (Exposure_group), need to be re-coded as dummy variables. Binary variables should be 0 or 1 (Government vs non-Government). In order to intemperate intercept of the model here we also center the numeric variables at the mean value and year at the start of the cohort (2008). Finally, interaction effects need to be created prior to the modelling. Grade 7 and 9 were combined due to relatively smaller numbers. Also the estimated effect size were also very similar when included separately.

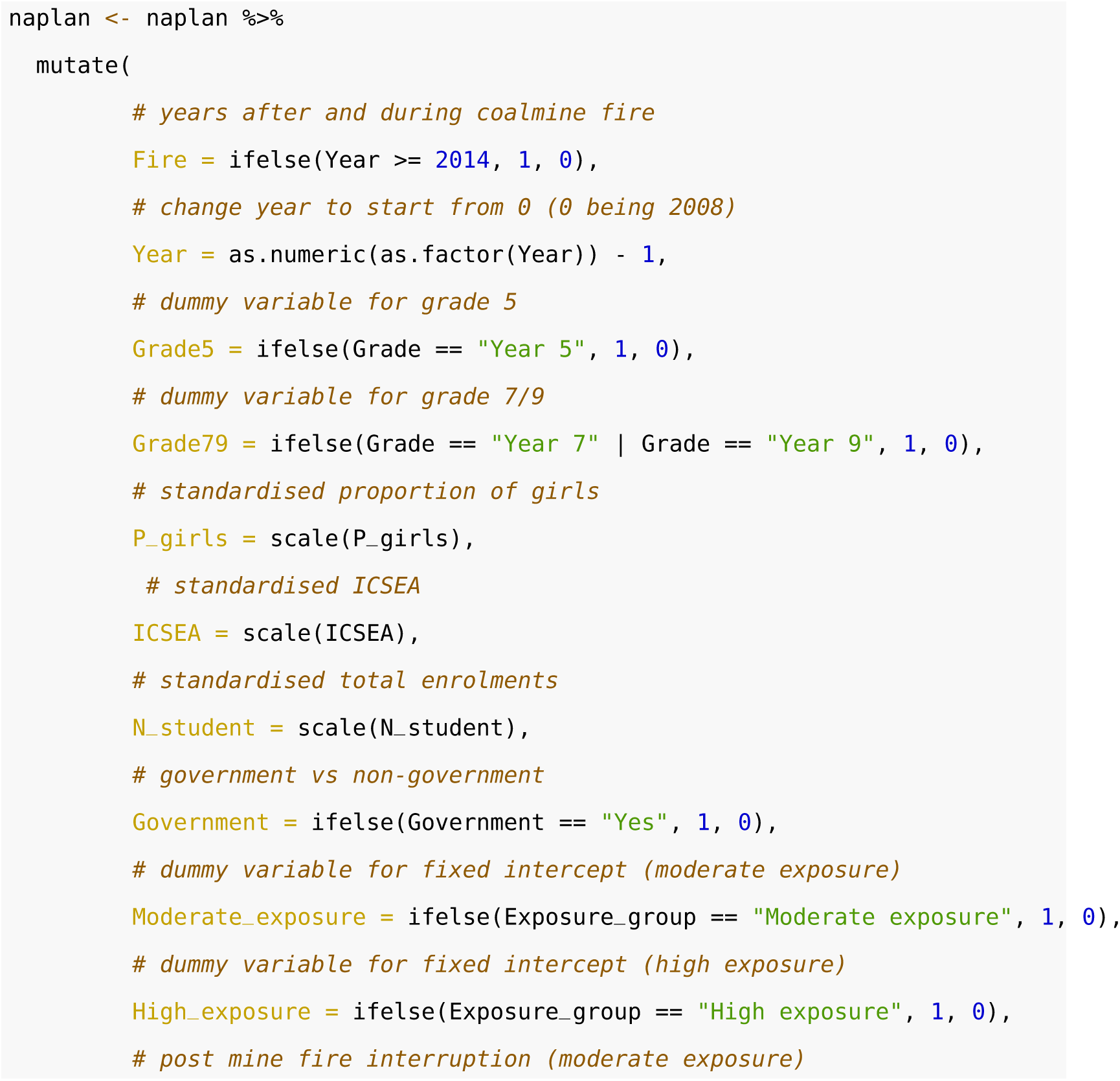

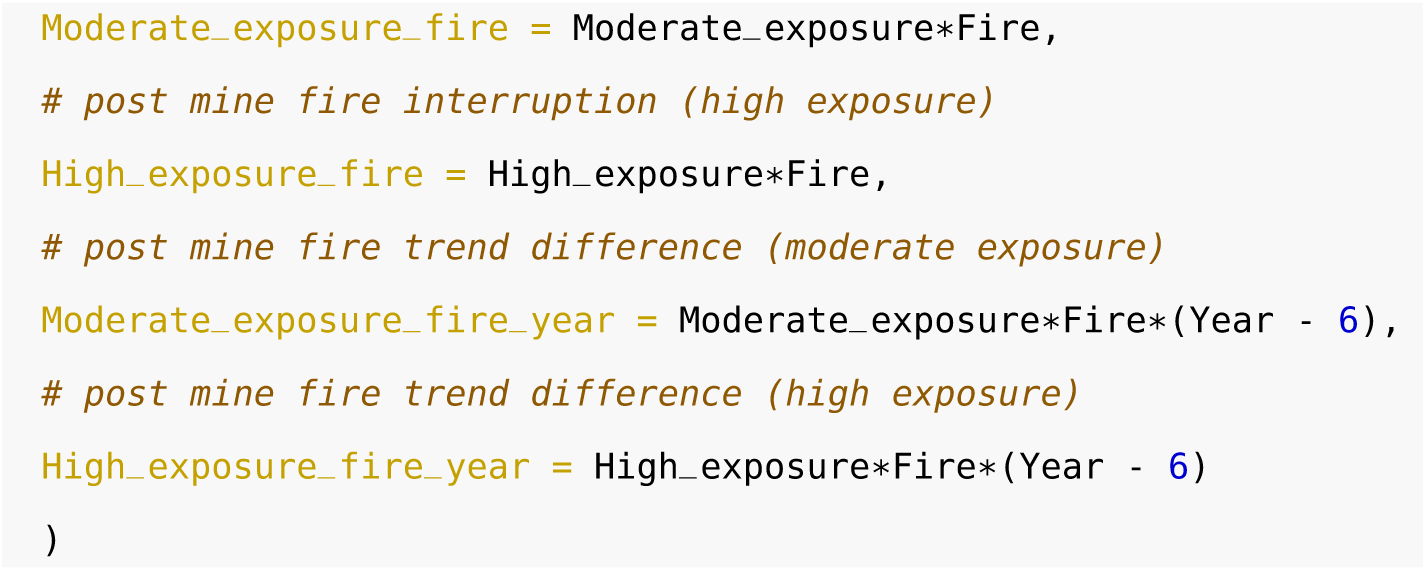

#### Stan model block

When using Rmarkdown file, stan code can be directly included as a block of code with specification of {stan output.var = “StanModel”} in the code block. In this model we use weakly informative priors, *N*(10, 5), for the SDs of the random school effects, random cohort effects as well as random error *N*(10, 5). 10 was chosen because when using two-level mixed-effects models with the mean score differences as the outcome variable, the estimated error terms are close to 10.

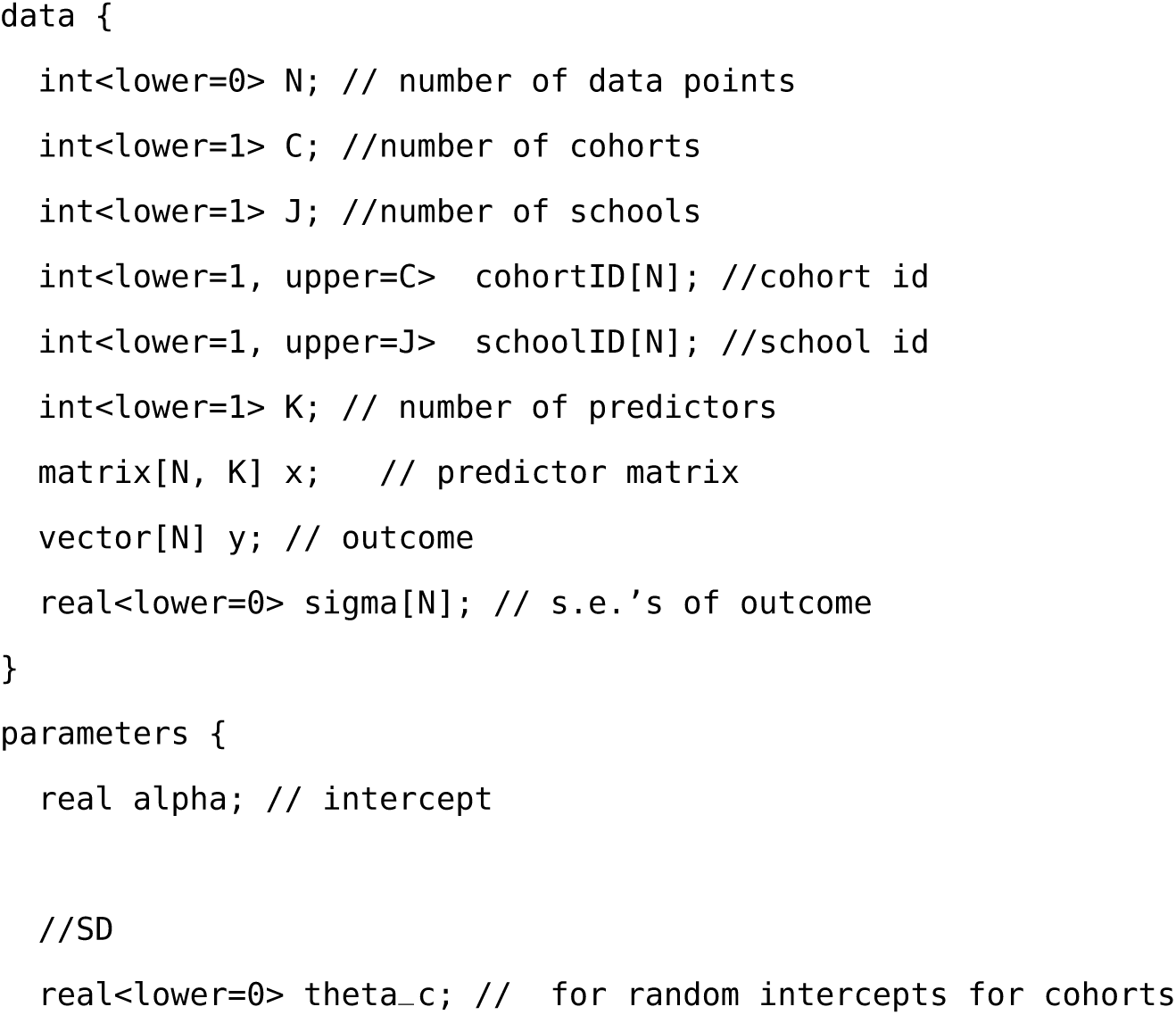

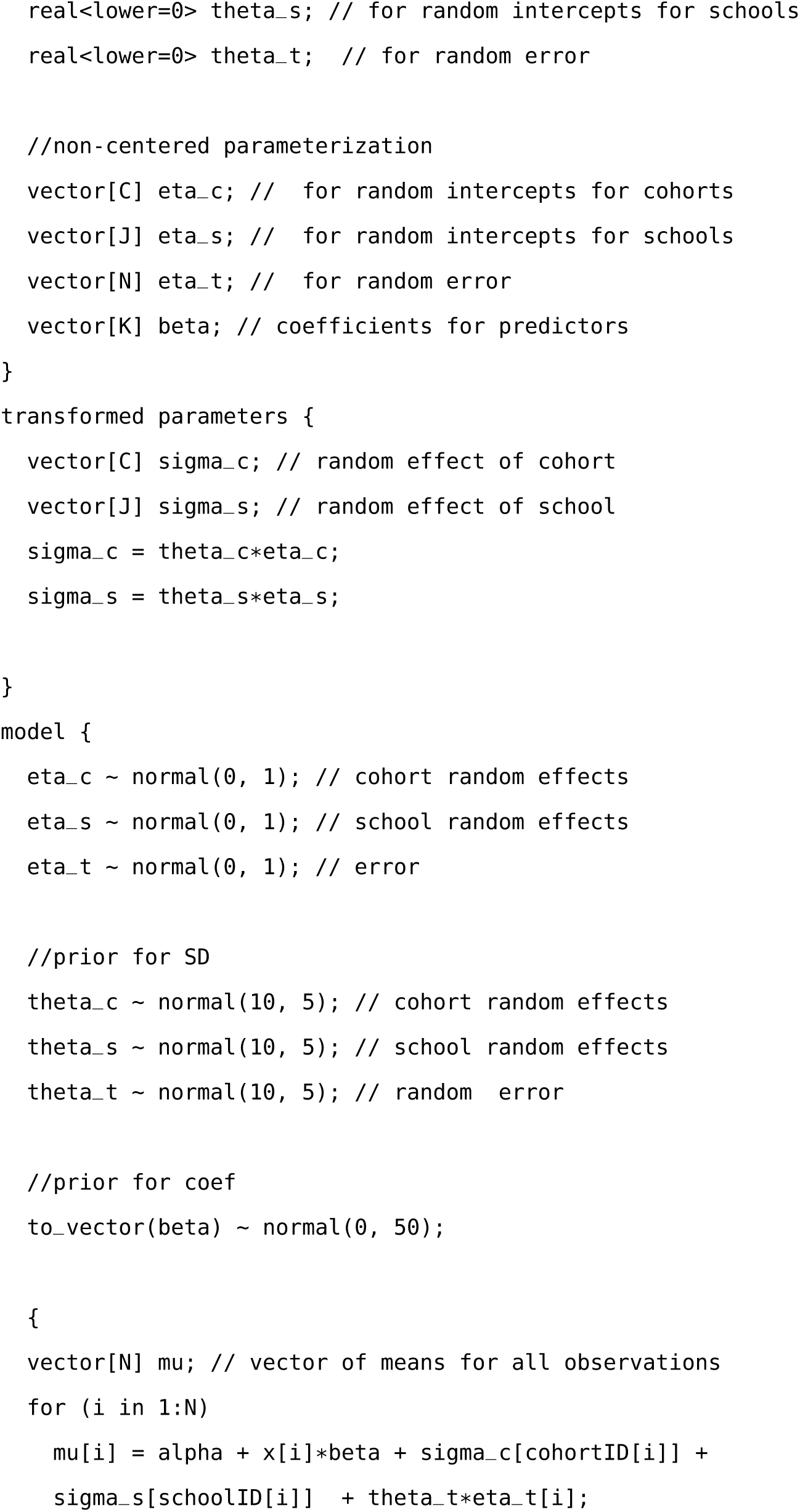

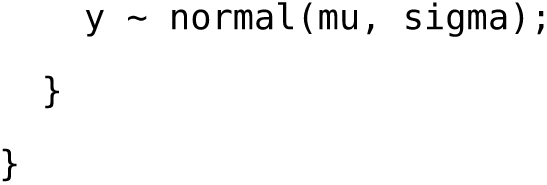

#### Run Stan model

Input data is needed to be saved in a list.

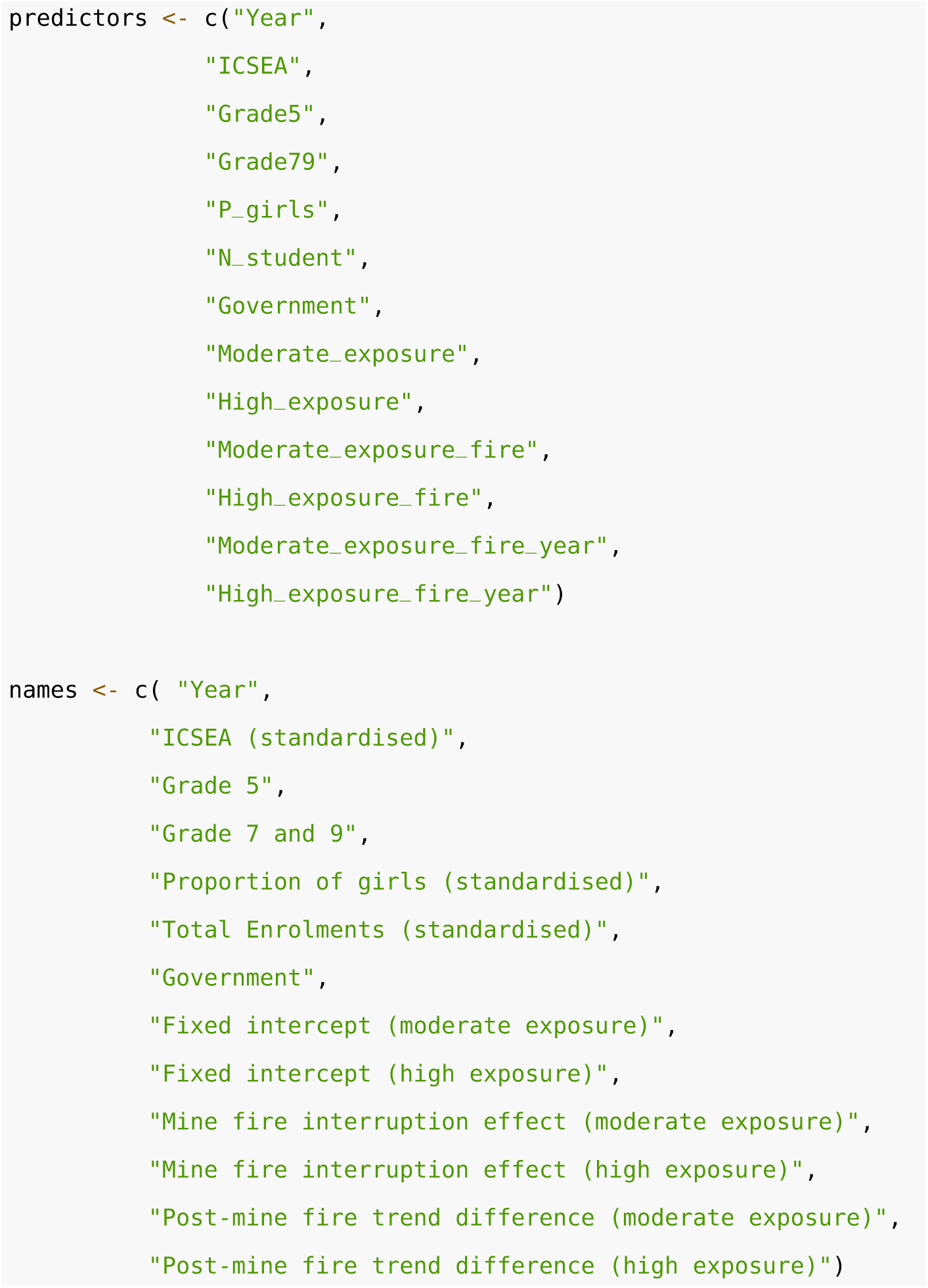

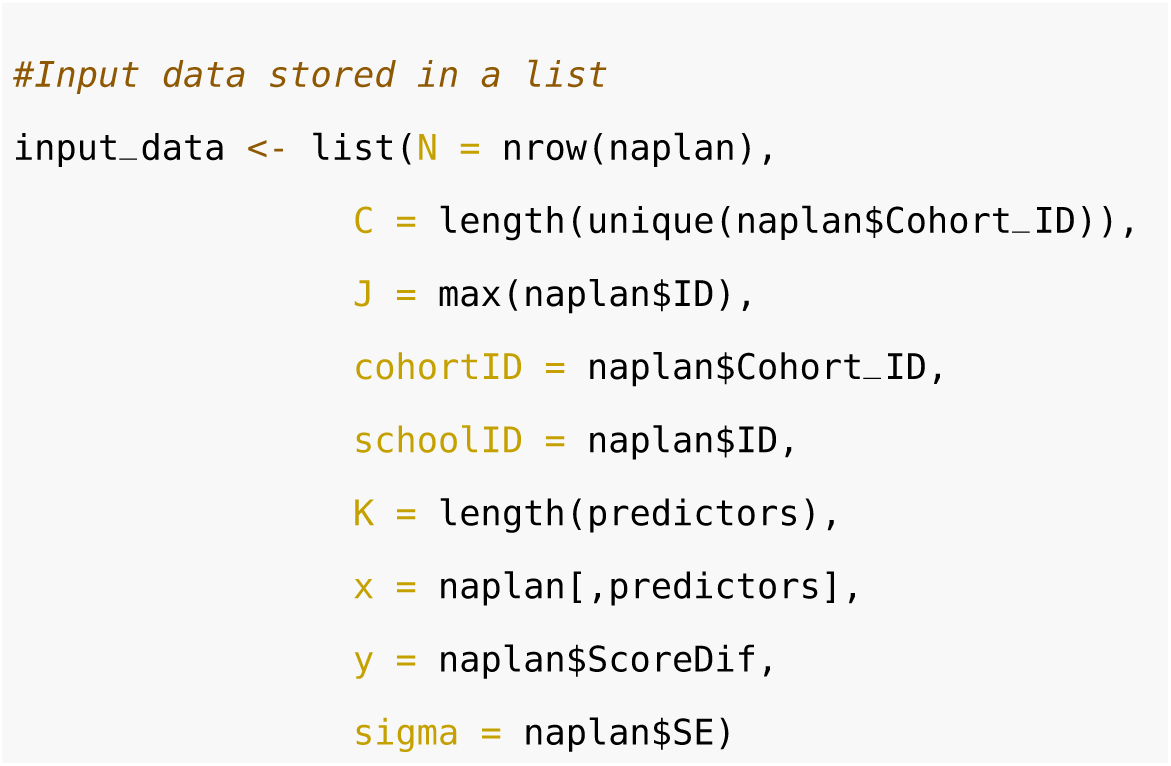

The next stage is to run the simulation using the Stan model defined as ‘StanModel’

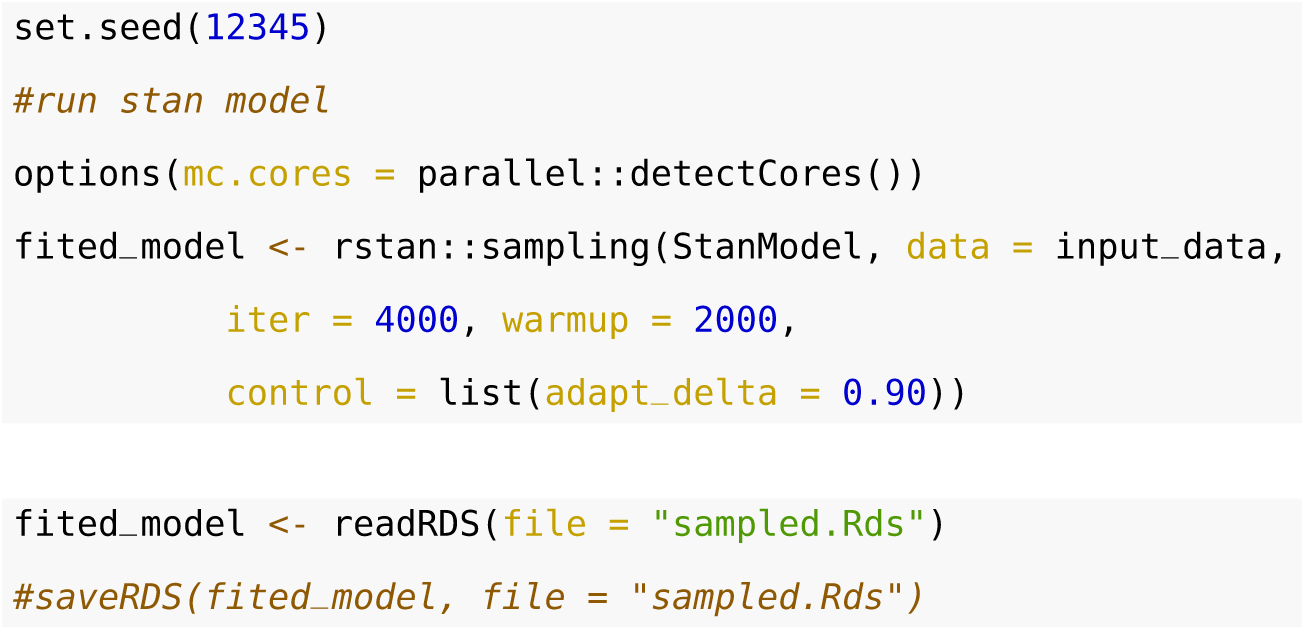

Next we extract results

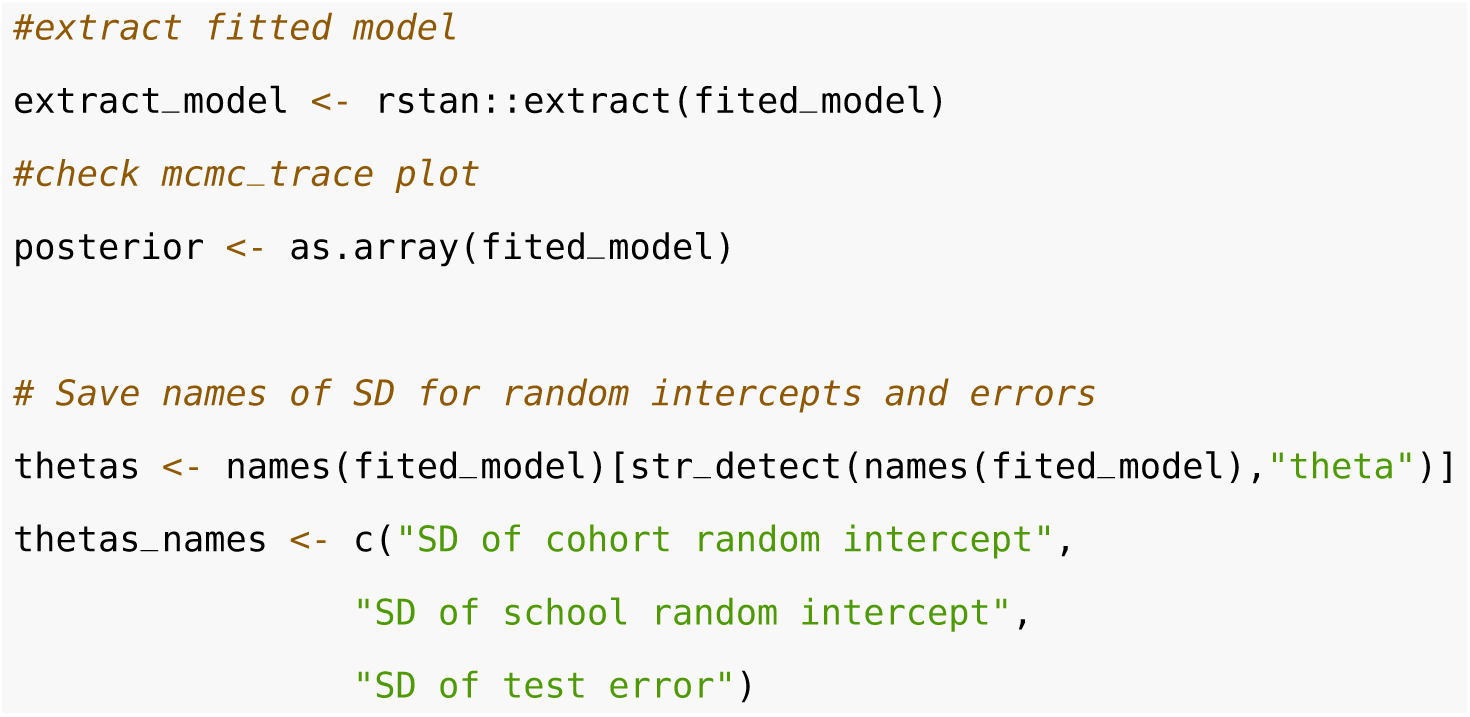

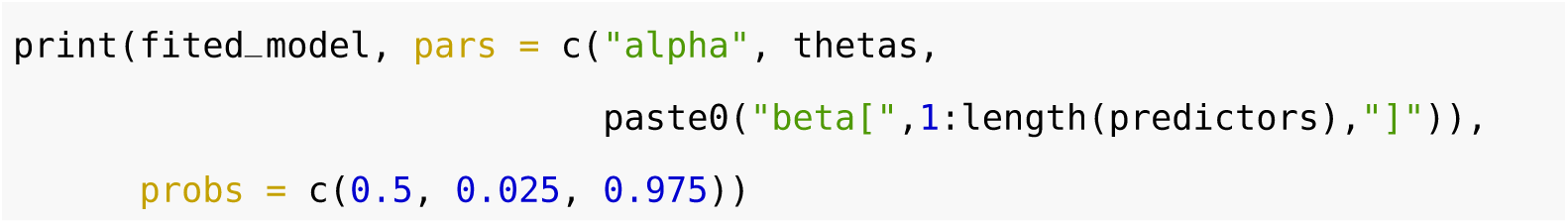

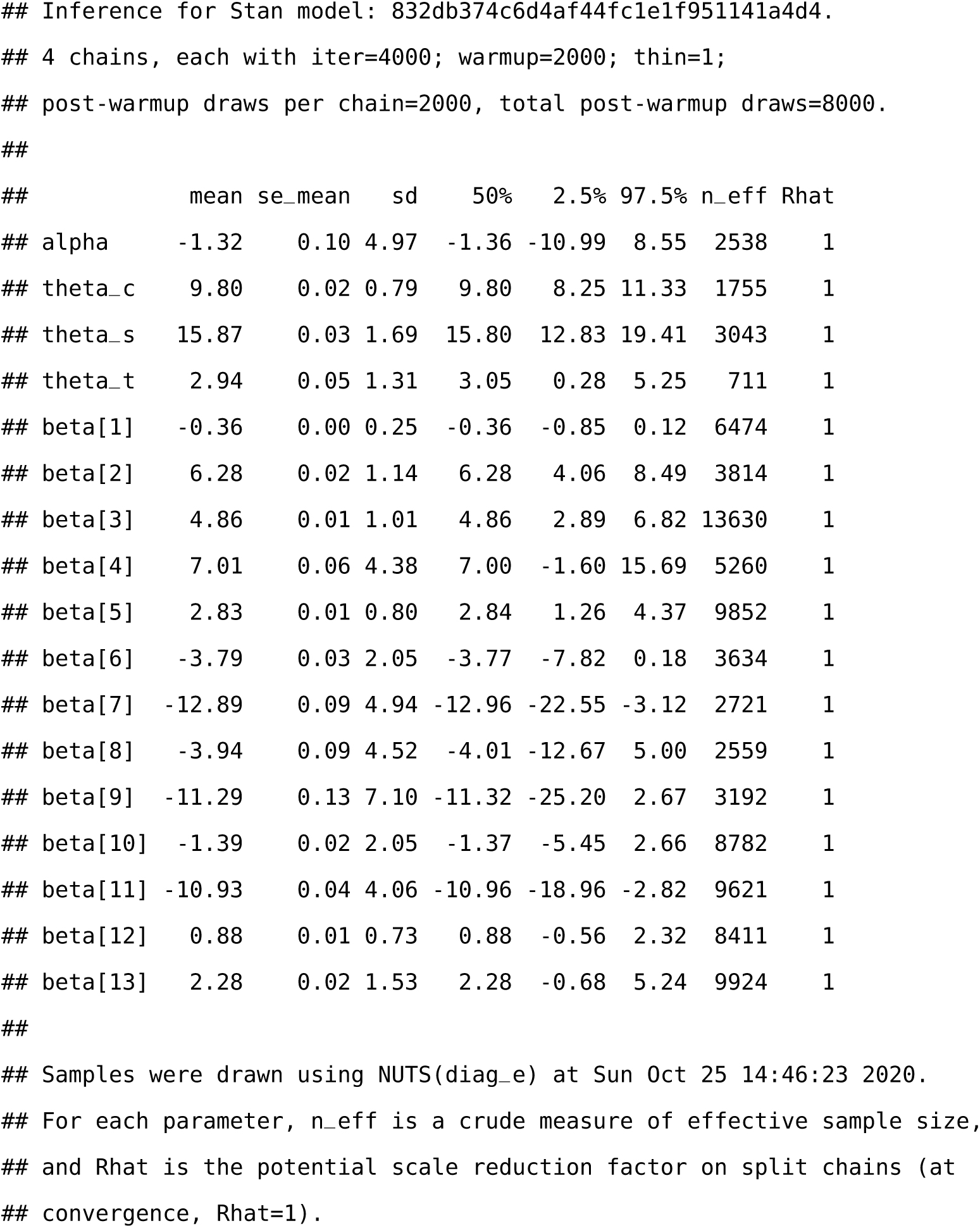

Full model diagnostics can be evaluated using an interactive shinystan package. Here we provide a few static diagnostic plots.

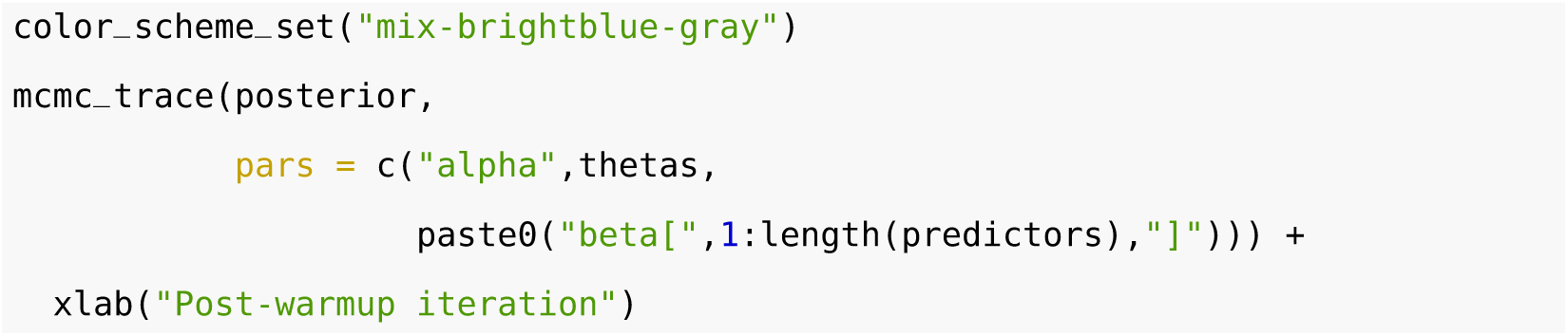

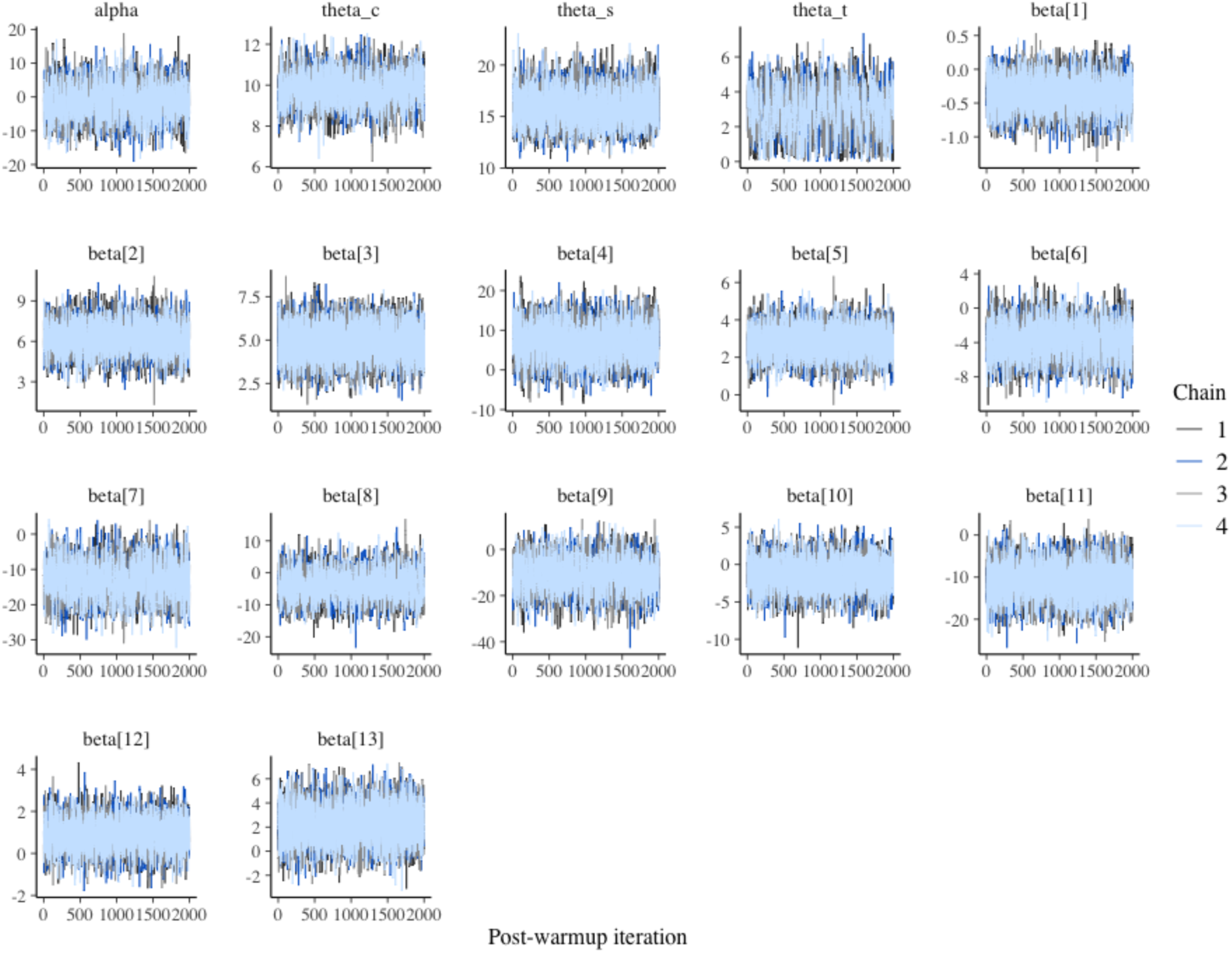

Markov chain Monte Carlo (MCMC) plot shows no signs of poor mixing for each coefficient. There was no warning of divergent transitions (using non-centred parameterisation centered parameterisation can help with avoiding divergent transitions), which can be diagnosed using diagnostic plots for the NUTS. All continuous variables are confounding variables (we are not interested in estimating effect sizes from these parameters), hence they were all standardised in the analysis to improve model fitting speed. If any variable of interest is a continuous variable, the original parameters can be easily recovered (see Stan manual) post standardisation.

The *mcmc_pairs* function is used to visualize the univariate histograms as well as bivariate scatter plots for key parameters. It is useful in identifying multicollinearity (strong correlation) and other non-identifiability issues (banana-like shapes).

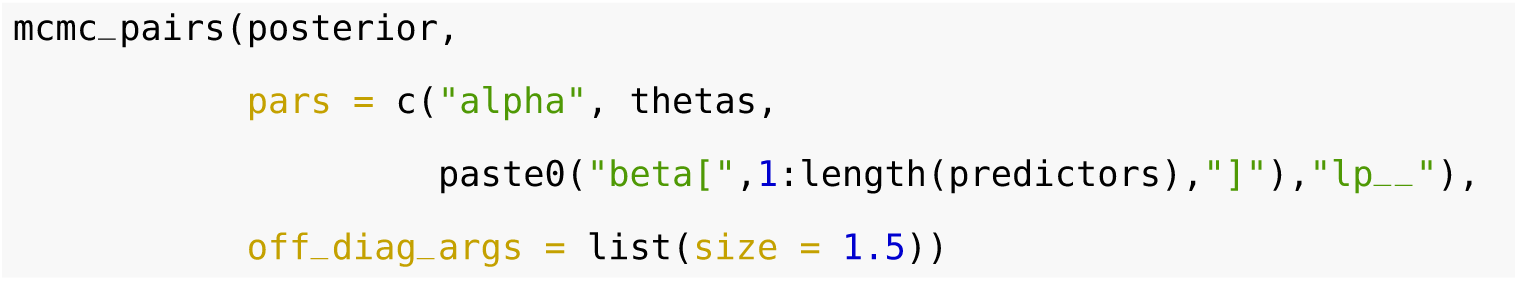

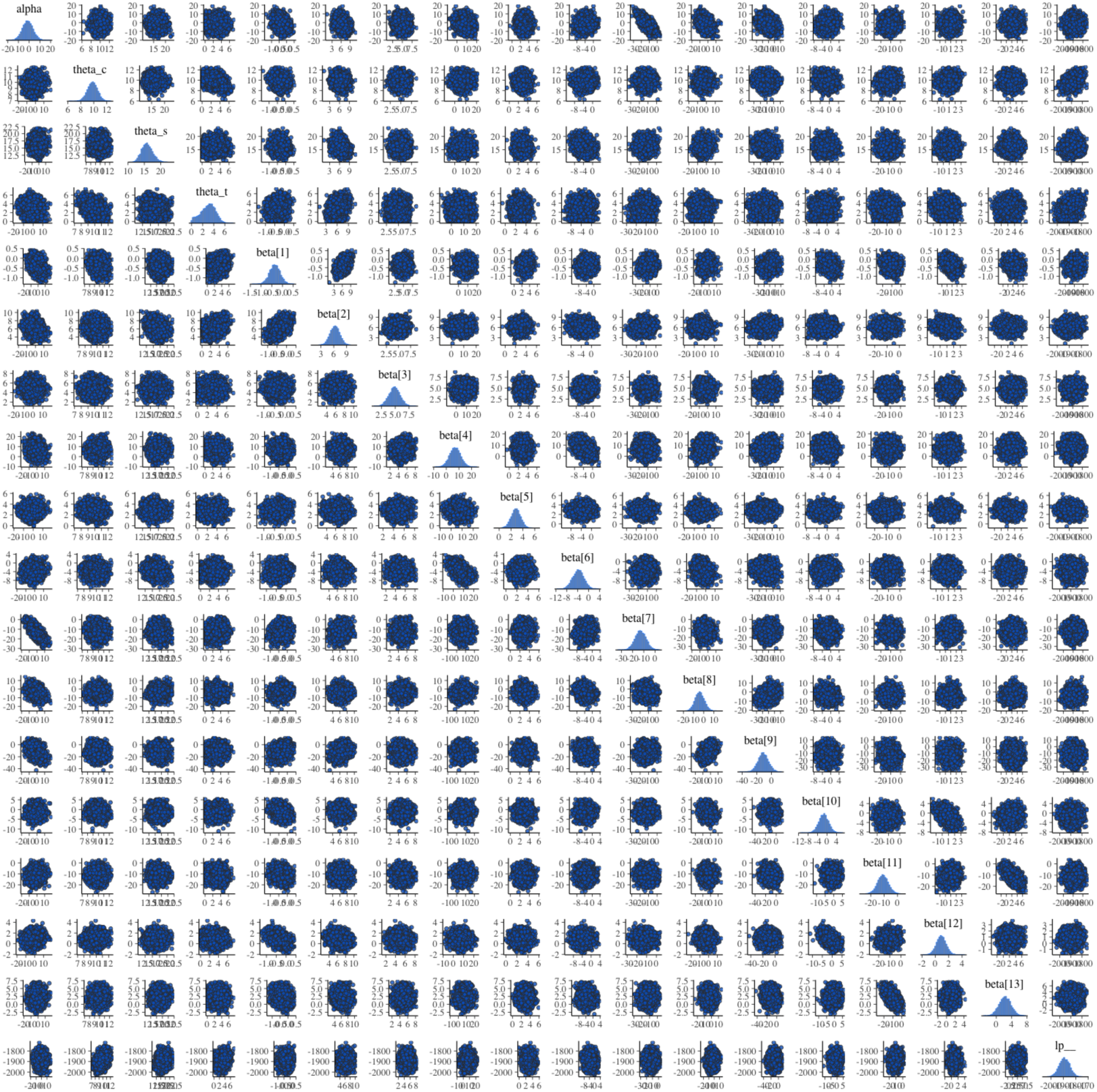

There is a negative association between sampled coefficients of the intercept term (alpha) and school sector (government, beta[7]). This is possible as school sector (government vs non-government) is the most important predictor of school-level NAPLAN results. Hence the sampled intercept will be impacted by the sampled coefficient of the school sector.

### Model summary

Mean and credible intervals

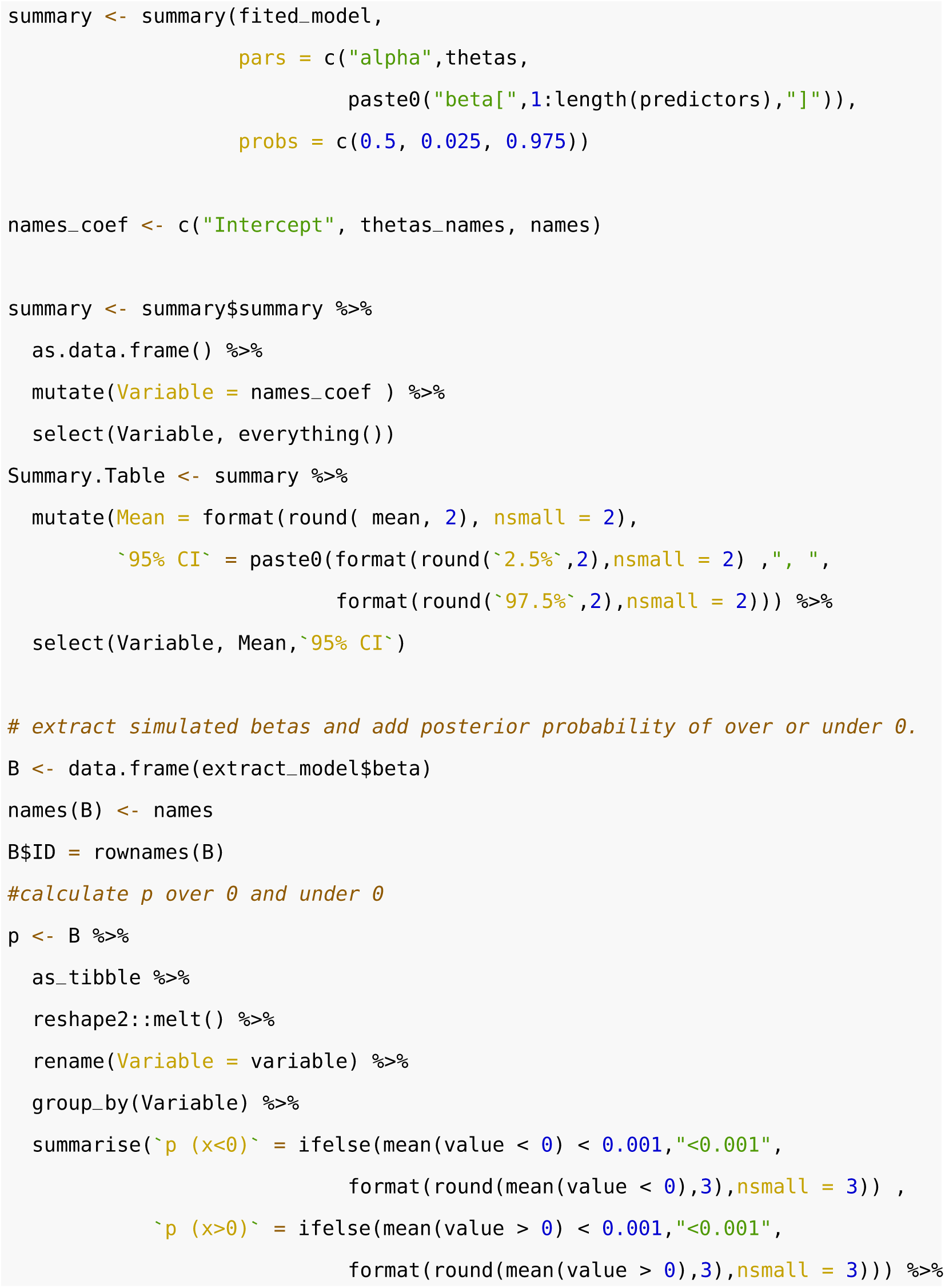

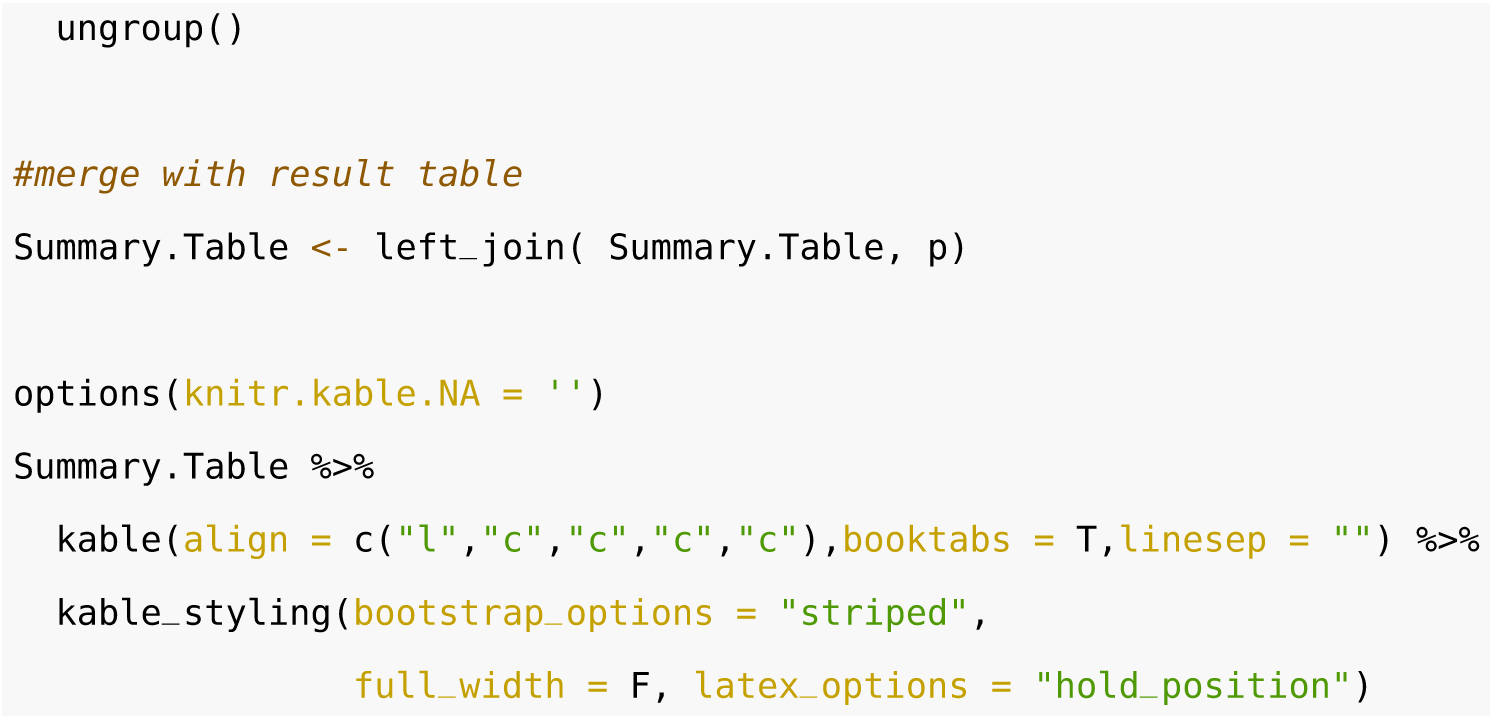

| Variable | Mean | 95% CI | p (x<0) | p (x>0) |
| --- | --- | --- | --- | --- |
| Intercept | -1.32 | -10.99, 8.55 |  |  |
| SD of cohort random intercept | 9.80 | 8.25, 11.33 |  |  |
| SD of school random intercept | 15.87 | 12.83, 19.41 |  |  |
| SD of test error | 2.94 | 0.28, 5.25 |  |  |
| Year | -0.36 | -0.85, 0.12 | 0.926 | 0.074 |
| ICSEA (standardised) | 6.28 | 4.06, 8.49 | <0.001 | 1.000 |
| Grade 5 | 4.86 | 2.89, 6.82 | <0.001 | 1.000 |
| Grade 7 and 9 | 7.01 | -1.60, 15.69 | 0.054 | 0.946 |
| Proportion of girls (standardised) | 2.83 | 1.26, 4.37 | <0.001 | 1.000 |
| Total Enrolments (standardised) | -3.79 | -7.82, 0.18 | 0.969 | 0.031 |
| Government | -12.89 | -22.55, -3.12 | 0.994 | 0.006 |
| Fixed intercept (moderate exposure) | -3.94 | -12.67, 5.00 | 0.808 | 0.192 |
| Fixed intercept (high exposure) | -11.29 | -25.20, 2.67 | 0.942 | 0.058 |
| Mine fire interruption effect (moderate exposure) | -1.39 | -5.45, 2.66 | 0.755 | 0.245 |
| Mine fire interruption effect (high exposure) | -10.93 | -18.96, -2.82 | 0.997 | 0.004 |
| Post-mine fire trend difference (moderate exposure) | 0.88 | -0.56, 2.32 | 0.113 | 0.887 |
| Post-mine fire trend difference (high exposure) | 2.28 | -0.68, 5.24 | 0.071 | 0.929 |

### Plot marginal effects

Here we obtain predicted margins using the posterior distribution of coefficients using the following steps:

- Obtain design matrix with confounding variables fixed at reference values
- Calculate posterior distribution of NAPLAN score difference by year and exposure zone
- Plot posterior mean (with error bar) of NAPLAN score difference for each year and exposure zone

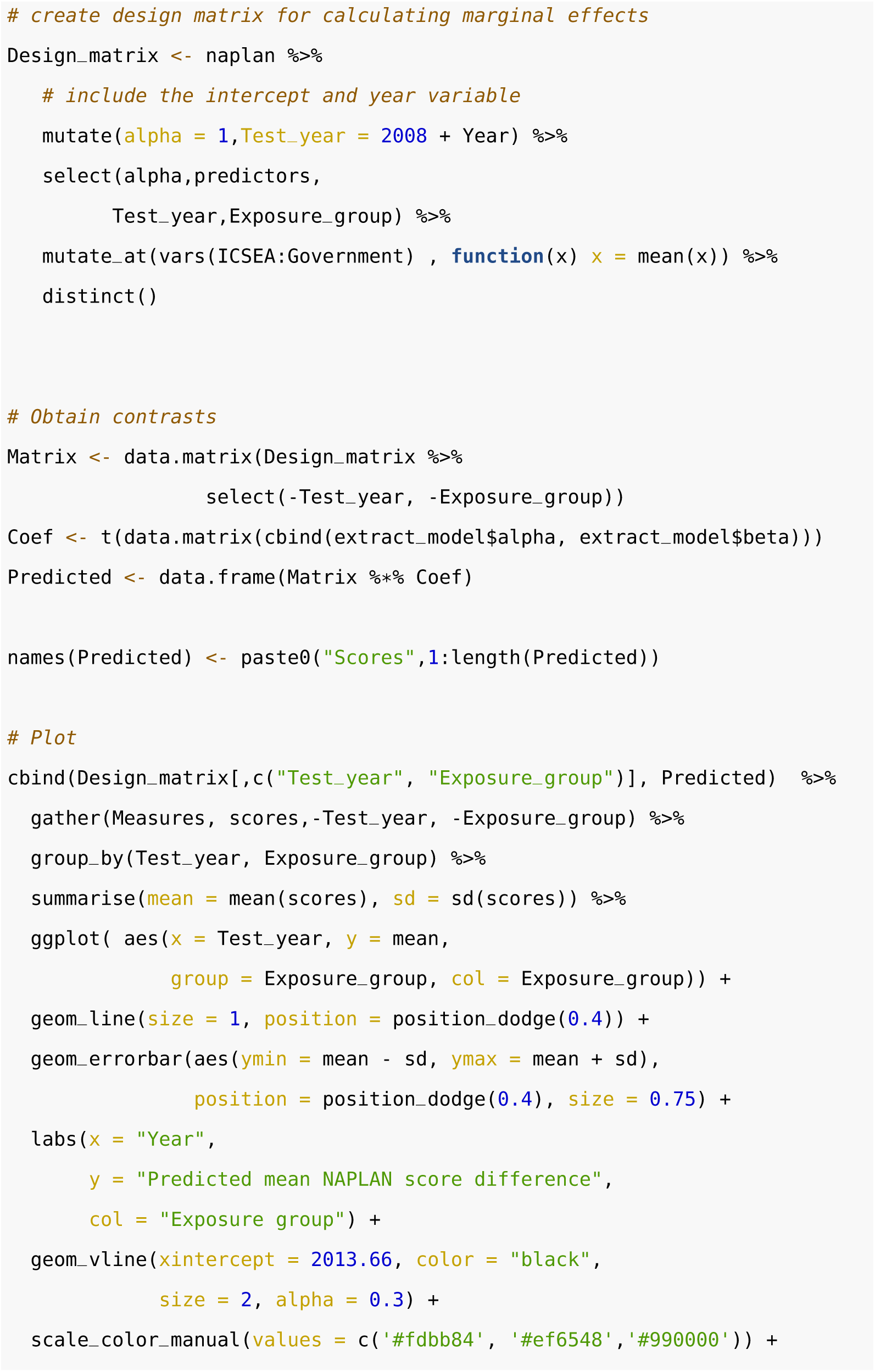

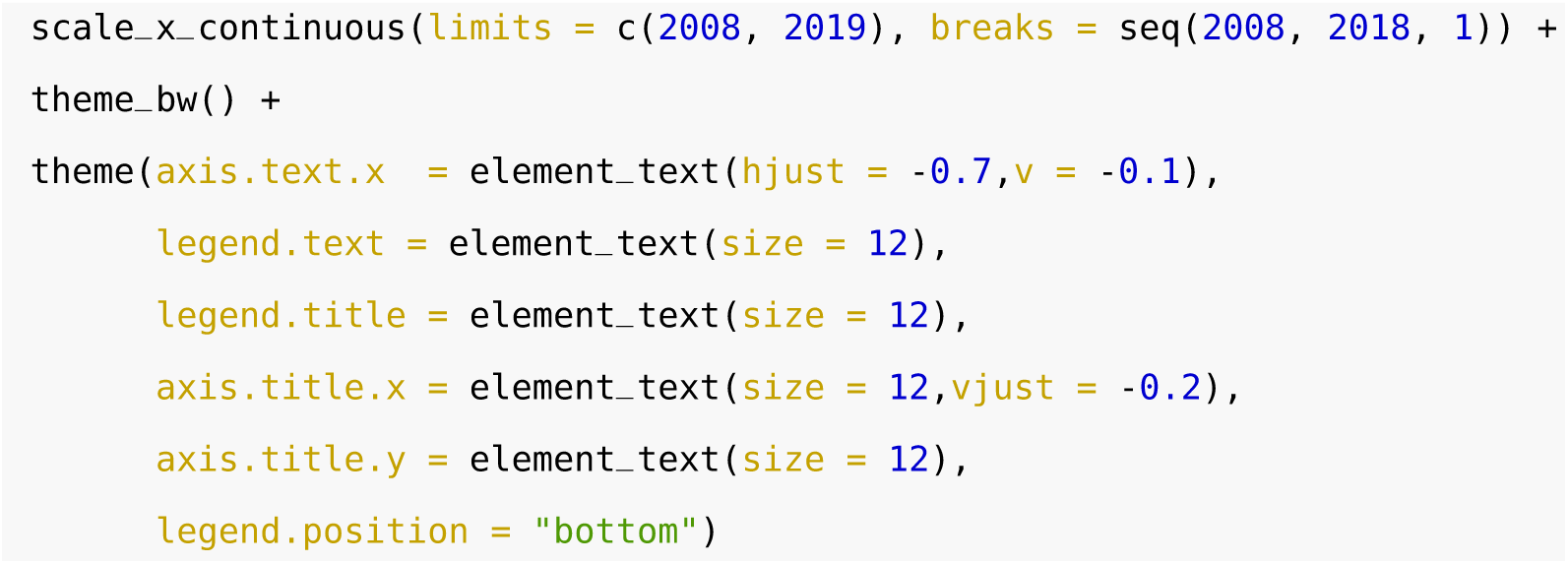

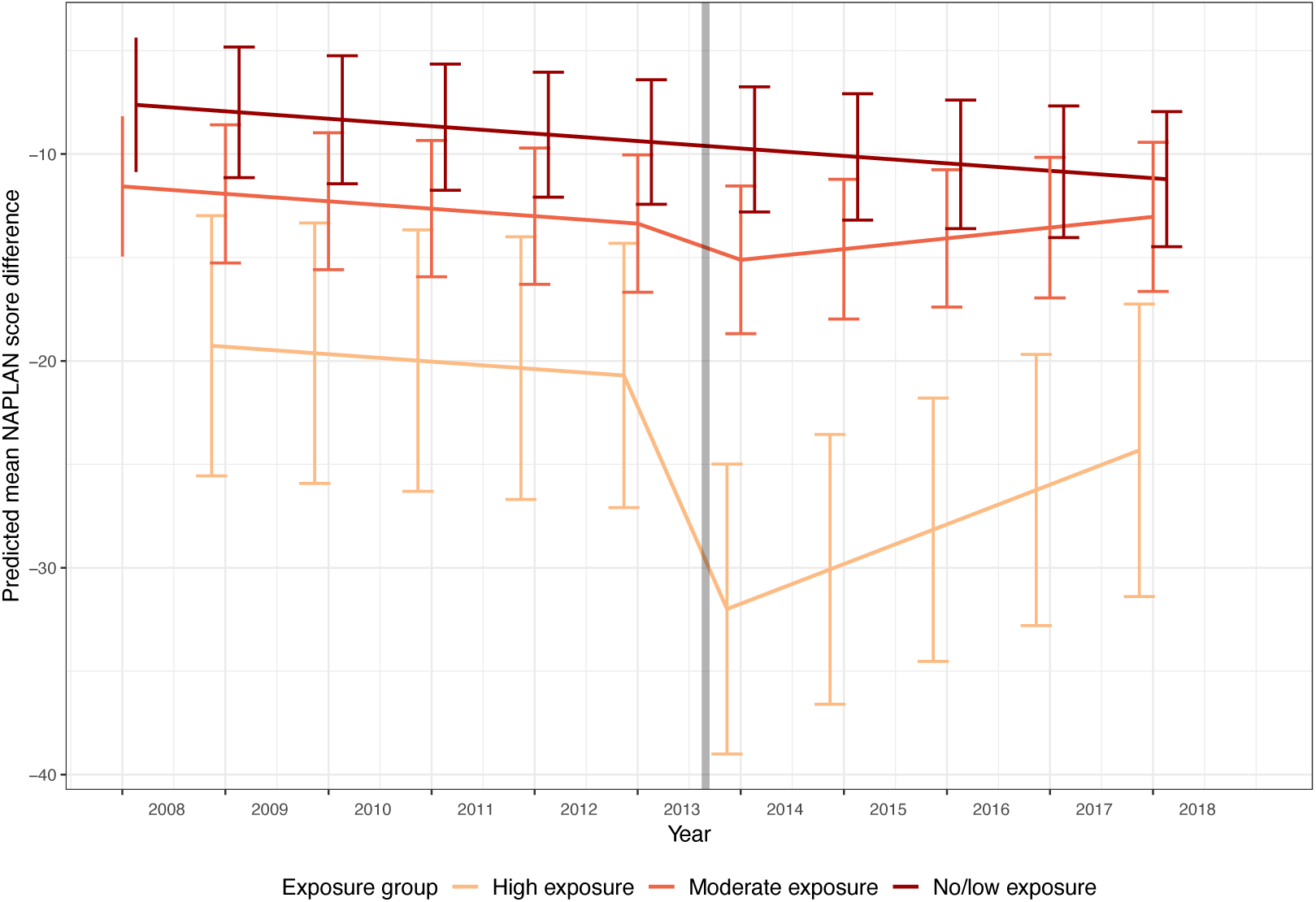

## Supplementary material II

**Table S1:** Bayesian hierarchical meta-regression models for grammar and punctuation

| | $\beta$ | 95% CI | $P(\beta < 0)$ | $P(\beta > 0)$ |
| --- | --- | --- | --- | --- |
| Intercept | -1.19 | -11.15, 8.74 |  |  |
| SD of cohort random intercept | 9.71 | 8.21, 11.25 |  |  |
| SD of school random intercept | 15.98 | 12.92, 19.49 |  |  |
| SD of test error | 2.69 | 0.22, 5.03 |  |  |
| Year | -0.37 | -0.85, 0.12 |  | 0.07 |
| ICSEA (standardised) | 6.12 | 3.91, 8.30 | <0.001 |  |
| Grade 5 | 4.73 | 2.69, 6.77 | <0.001 |  |
| Grade 7 and 9 | 7.22 | -1.15, 15.68 | 0.048 |  |
| Proportion of girls (standardised) | 2.81 | 1.21, 4.39 | <0.001 |  |
| Total enrolments (standardised) | -3.79 | -7.73, 0.13 |  | 0.029 |
| Government | -12.98 | -22.62, -3.34 |  | 0.003 |
| Fixed intercept (moderate exposure) | -3.90 | -13.13, 5.31 |  | 0.20 |
| Fixed intercept (high exposure) | -11.61 | -25.28, 2.18 |  | 0.05 |
| Mine fire interruption effect (moderate exposure) | -1.78 | -5.73, 2.13 |  | 0.19 |
| Mine fire interruption effect (high exposure) | -10.91 | -18.68, -3.08 |  | 0.004 |
| Post-mine fire trend difference (moderate exposure) | 0.92 | -0.51, 2.35 | 0.11 |  |
| Post-mine fire trend difference (high exposure) | 2.37 | -0.73, 5.37 | 0.06 |  |

**Table S2:** Bayesian hierarchical meta-regression models for numeracy

| | $\beta$ | 95% CI | $P(\beta < 0)$ | $P(\beta > 0)$ |
| --- | --- | --- | --- | --- |
| Intercept | -2.09 | -11.43, 7.13 |  |  |
| SD of cohort random intercept | 9.71 | 8.34, 11.06 |  |  |
| SD of school random intercept | 14.67 | 11.84, 17.86 |  |  |
| SD of test error | 5.66 | 4.17, 7.12 |  |  |
| Year | -0.38 | -0.84, 0.07 |  | 0.05 |
| ICSEA (standardised) | 5.98 | 3.95, 8.00 | <0.001 |  |
| Grade 5 | 3.40 | 1.69, 5.12 | <0.001 |  |
| Grade 7 and 9 | 11.85 | 3.60, 20.18 | 0.004 |  |
| Proportion of girls (standardised) | 1.14 | -0.25, 2.52 | 0.05 |  |
| Total enrolments (standardised) | -4.75 | -8.43, -0.93 |  | 0.008 |
| Government | -7.93 | -17.02, 0.92 |  | 0.042 |
| Fixed intercept (moderate exposure) | -5.66 | -13.94, 2.60 |  | 0.08 |
| Fixed intercept (high exposure) | -12.07 | -24.26, 0.23 |  | 0.028 |
| Mine fire interruption effect (moderate exposure) | -0.44 | -4.20, 3.28 |  | 0.41 |
| Mine fire interruption effect (high exposure) | -10.90 | -17.98, -3.67 |  | <0.001 |
| Post-mine fire trend difference (moderate exposure) | 0.79 | -0.56, 2.13 | 0.13 |  |
| Post-mine fire trend difference (high exposure) | 1.61 | -1.21, 4.43 | 0.13 |  |

**Table S3:** Bayesian hierarchical meta-regression models for reading

| | $\beta$ | 95% CI | $P(\beta < 0)$ | $P(\beta > 0)$ |
| --- | --- | --- | --- | --- |
| Intercept | -3.70 | -13.03, 5.71 |  |  |
| SD of cohort random intercept | 9.47 | 8.05, 10.90 |  |  |
| SD of school random intercept | 15.05 | 12.05, 18.55 |  |  |
| SD of test error | 2.78 | 0.48, 4.65 |  |  |
| Year | -0.37 | -0.82, 0.10 |  | 0.06 |
| ICSEA (standardised) | 5.83 | 3.84, 7.93 | <0.001 |  |
| Grade 5 | 3.77 | 1.90, 5.71 | <0.001 |  |
| Grade 7 and 9 | 7.66 | -0.35, 15.70 | 0.030 |  |
| Proportion of girls (standardised) | 1.93 | 0.48, 3.41 | 0.005 |  |
| Total enrolments (standardised) | -3.77 | -7.62, -0.08 |  | 0.022 |
| Government | -9.77 | -19.02, -0.30 |  | 0.022 |
| Fixed intercept (moderate exposure) | -1.87 | -10.19, 6.39 |  | 0.33 |
| Fixed intercept (high exposure) | -13.41 | -26.26, -0.19 |  | 0.024 |
| Mine fire interruption effect (moderate exposure) | -1.18 | -4.84, 2.57 |  | 0.27 |
| Mine fire interruption effect (high exposure) | -8.34 | -15.51, -1.07 |  | 0.013 |
| Post-mine fire trend difference (moderate exposure) | 0.69 | -0.61, 2.01 | 0.15 |  |
| Post-mine fire trend difference (high exposure) | 1.05 | -1.63, 3.85 | 0.23 |  |

**Table S4:** Bayesian hierarchical meta-regression models for spelling

| | $\beta$ | 95% CI | $P(\beta < 0)$ | $P(\beta > 0)$ |
| --- | --- | --- | --- | --- |
| Intercept | -0.79 | -9.41, 7.90 |  |  |
| SD of cohort random intercept | 10.38 | 9.19, 11.59 |  |  |
| SD of school random intercept | 13.64 | 11.05, 16.70 |  |  |
| SD of test error | 1.31 | 0.07, 3.04 |  |  |
| Year | -0.48 | -0.91, -0.03 |  | 0.016 |
| ICSEA (standardised) | 4.54 | 2.63, 6.41 | <0.001 |  |
| Grade 5 | 3.62 | 1.87, 5.37 | <0.001 |  |
| Grade 7 and 9 | 5.35 | -2.65, 13.79 | 0.10 |  |
| Proportion of girls (standardised) | 1.38 | -0.07, 2.82 | 0.030 |  |
| Total enrolments (standardised) | -0.94 | -4.61, 2.74 |  | 0.31 |
| Government | -8.05 | -16.52, 0.07 |  | 0.026 |
| Fixed intercept (moderate exposure) | -0.52 | -8.45, 7.05 |  | 0.45 |
| Fixed intercept (high exposure) | -3.68 | -16.16, 8.34 |  | 0.27 |
| Mine fire interruption effect (moderate exposure) | -1.44 | -4.91, 2.09 |  | 0.21 |
| Mine fire interruption effect (high exposure) | -10.31 | -17.39, -3.38 |  | 0.003 |
| Post-mine fire trend difference (moderate exposure) | -0.42 | -1.68, 0.85 | 0.74 |  |
| Post-mine fire trend difference (high exposure) | -1.49 | -4.20, 1.21 | 0.86 |  |

**Table S5:** Bayesian hierarchical meta-regression models for writing

| | $\beta$ | 95% CI | $P(\beta < 0)$ | $P(\beta > 0)$ |
| --- | --- | --- | --- | --- |
| Intercept | -1.50 | -10.28, 7.31 |  |  |
| SD of cohort random intercept | 6.38 | 4.50, 8.03 |  |  |
| SD of school random intercept | 13.94 | 11.41, 16.87 |  |  |
| SD of test error | 7.80 | 6.36, 9.20 |  |  |
| Year | -0.62 | -1.06, -0.16 |  | 0.002 |
| ICSEA (standardised) | 8.66 | 6.64, 10.66 | <0.001 |  |
| Grade 5 | 3.26 | 1.35, 5.15 | <0.001 |  |
| Grade 7 and 9 | 12.12 | 4.43, 19.72 | <0.001 |  |
| Proportion of girls (standardised) | 1.57 | 0.19, 3.00 | 0.012 |  |
| Total enrolments (standardised) | -4.70 | -8.38, -0.97 |  | 0.006 |
| Government | -13.89 | -22.57, -5.43 |  | <0.001 |
| Fixed intercept (moderate exposure) | 1.01 | -6.83, 8.71 |  | 0.61 |
| Fixed intercept (high exposure) | -7.53 | -19.63, 4.62 |  | 0.11 |
| Mine fire interruption effect (moderate exposure) | 3.56 | -0.52, 7.55 |  | 0.96 |
| Mine fire interruption effect (high exposure) | -11.09 | -18.93, -3.16 |  | 0.003 |
| Post-mine fire trend difference (moderate exposure) | -0.60 | -2.01, 0.82 | 0.80 |  |
| Post-mine fire trend difference (high exposure) | 2.44 | -0.60, 5.50 | 0.06 |  |

**Table S6:**
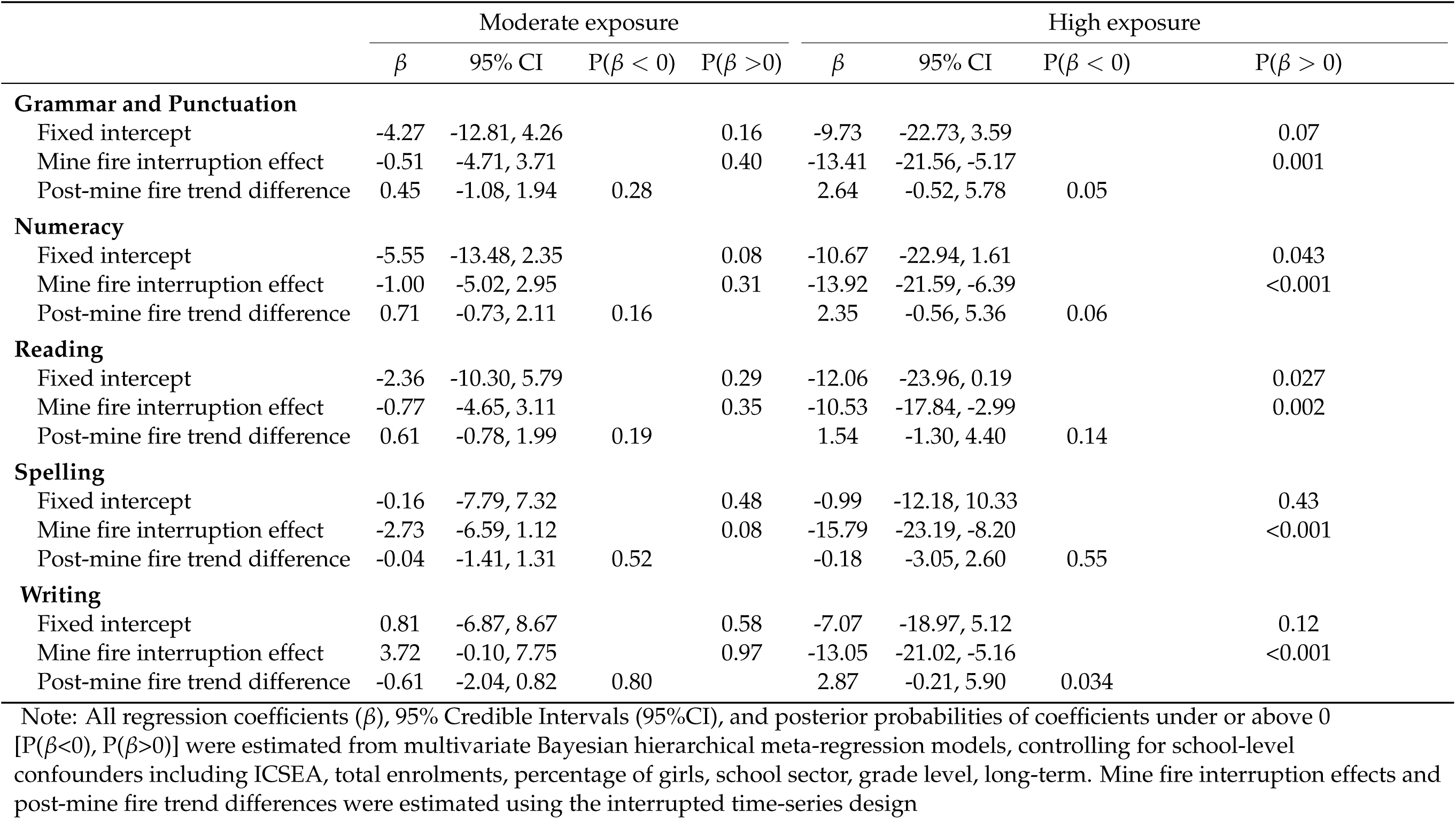
Estimated intercept, mine fire interruption effect and post-mine fire trend difference for moderate and high exposure schools estimated from Bayesian hierarchical meta-regression models (excluding cohort effect)

|  | Moderate exposure |  |  |  | High exposure |  |  |  |
| --- | --- | --- | --- | --- | --- | --- | --- | --- |
| | $\beta$ | 95% CI | $P(\beta < 0)$ | $P(\beta > 0)$ | $\beta$ | 95% CI | $P(\beta < 0)$ | $P(\beta > 0)$ |
| **Grammar and Punctuation** |  |  |  |  |  |  |  |  |
| Fixed intercept | -4.27 | -12.81, 4.26 |  | 0.16 | -9.73 | -22.73, 3.59 |  | 0.07 |
| Mine fire interruption effect | -0.51 | -4.71, 3.71 |  | 0.40 | -13.41 | -21.56, -5.17 |  | 0.001 |
| Post-mine fire trend difference | 0.45 | -1.08, 1.94 | 0.28 |  | 2.64 | -0.52, 5.78 | 0.05 |  |
| **Numeracy** |  |  |  |  |  |  |  |  |
| Fixed intercept | -5.55 | -13.48, 2.35 |  | 0.08 | -10.67 | -22.94, 1.61 |  | 0.043 |
| Mine fire interruption effect | -1.00 | -5.02, 2.95 |  | 0.31 | -13.92 | -21.59, -6.39 |  | <0.001 |
| Post-mine fire trend difference | 0.71 | -0.73, 2.11 | 0.16 |  | 2.35 | -0.56, 5.36 | 0.06 |  |
| **Reading** |  |  |  |  |  |  |  |  |
| Fixed intercept | -2.36 | -10.30, 5.79 |  | 0.29 | -12.06 | -23.96, 0.19 |  | 0.027 |
| Mine fire interruption effect | -0.77 | -4.65, 3.11 |  | 0.35 | -10.53 | -17.84, -2.99 |  | 0.002 |
| Post-mine fire trend difference | 0.61 | -0.78, 1.99 | 0.19 |  | 1.54 | -1.30, 4.40 | 0.14 |  |
| **Spelling** |  |  |  |  |  |  |  |  |
| Fixed intercept | -0.16 | -7.79, 7.32 |  | 0.48 | -0.99 | -12.18, 10.33 |  | 0.43 |
| Mine fire interruption effect | -2.73 | -6.59, 1.12 |  | 0.08 | -15.79 | -23.19, -8.20 |  | <0.001 |
| Post-mine fire trend difference | -0.04 | -1.41, 1.31 | 0.52 |  | -0.18 | -3.05, 2.60 | 0.55 |  |
| **Writing** |  |  |  |  |  |  |  |  |
| Fixed intercept | 0.81 | -6.87, 8.67 |  | 0.58 | -7.07 | -18.97, 5.12 |  | 0.12 |
| Mine fire interruption effect | 3.72 | -0.10, 7.75 |  | 0.97 | -13.05 | -21.02, -5.16 |  | <0.001 |
| Post-mine fire trend difference | -0.61 | -2.04, 0.82 | 0.80 |  | 2.87 | -0.21, 5.90 | 0.034 |  |

Note: All regression coefficients ( $\beta$ ), 95% Credible Intervals (95%CI), and posterior probabilities of coefficients under or above 0 [ $P(\beta < 0)$ , $P(\beta > 0)$ ] were estimated from multivariate Bayesian hierarchical meta-regression models, controlling for school-level confounders including ICSEA, total enrolments, percentage of girls, school sector, grade level, long-term. Mine fire interruption effects and post-mine fire trend differences were estimated using the interrupted time-series design

**Table S7:** Estimated intercept, mine fire interruption effect and post-mine fire trend difference for moderate and high exposure schools estimated from Bayesian hierarchical meta-regression models (excluding relocated schools)

|  | Moderate exposure |  |  |  | High exposure |  |  |  |
| --- | --- | --- | --- | --- | --- | --- | --- | --- |
| | $\beta$ | 95% CI | $P(\beta < 0)$ | $P(\beta > 0)$ | $\beta$ | 95% CI | $P(\beta < 0)$ | $P(\beta > 0)$ |
| **Grammar and Punctuation** |  |  |  |  |  |  |  |  |
| Fixed intercept | -3.66 | -12.53, 5.03 |  | 0.21 | -12.03 | -27.29, 3.05 |  | 0.06 |
| Mine fire interruption effect | -1.96 | -5.93, 1.97 |  | 0.16 | -8.51 | -17.39, 0.23 |  | 0.027 |
| Post-mine fire trend difference | 0.87 | -0.53, 2.29 | 0.12 |  | 1.62 | -1.61, 4.80 | 0.16 |  |
| **Numeracy** |  |  |  |  |  |  |  |  |
| Fixed intercept | -5.06 | -13.33, 3.40 |  | 0.11 | -12.22 | -26.71, 1.56 |  | 0.041 |
| Mine fire interruption effect | -0.70 | -4.35, 3.05 |  | 0.35 | -7.56 | -15.76, 0.73 |  | 0.036 |
| Post-mine fire trend difference | 0.70 | -0.64, 2.01 | 0.15 |  | 1.08 | -1.95, 4.09 | 0.24 |  |
| **Reading** |  |  |  |  |  |  |  |  |
| Fixed intercept | -1.77 | -10.09, 6.51 |  | 0.34 | -15.64 | -30.60, -1.11 |  | 0.018 |
| Mine fire interruption effect | -1.28 | -4.88, 2.43 |  | 0.24 | -5.58 | -13.63, 2.43 |  | 0.08 |
| Post-mine fire trend difference | 0.64 | -0.67, 1.96 | 0.17 |  | 1.19 | -1.85, 4.19 | 0.22 |  |
| **Spelling** |  |  |  |  |  |  |  |  |
| Fixed intercept | -0.23 | -7.62, 7.54 |  | 0.46 | -5.96 | -19.44, 7.43 |  | 0.19 |
| Mine fire interruption effect | -1.65 | -5.14, 1.81 |  | 0.18 | -7.77 | -15.87, 0.05 |  | 0.026 |
| Post-mine fire trend difference | -0.47 | -1.72, 0.76 | 0.77 |  | -2.09 | -5.07, 0.73 | 0.92 |  |
| **Writing** |  |  |  |  |  |  |  |  |
| Fixed intercept | 1.25 | -6.57, 9.06 |  | 0.62 | -9.44 | -23.14, 4.23 |  | 0.08 |
| Mine fire interruption effect | 3.33 | -0.49, 7.25 |  | 0.96 | -9.95 | -18.76, -1.17 |  | 0.014 |
| Post-mine fire trend difference | -0.70 | -2.11, 0.72 | 0.84 |  | 2.30 | -1.00, 5.55 | 0.09 |  |

Note: All regression coefficients ( $\beta$ ), 95% Credible Intervals (95%CI), and posterior probabilities of coefficients under or above 0 [ $P(\beta < 0)$ , $P(\beta > 0)$ ] were estimated from multivariate Bayesian hierarchical meta-regression models, controlling for school-level confounders including ICSEA, total enrolments, percentage of girls, school sector, grade level, long-term. Mine fire interruption effects and post-mine fire trend differences were estimated using the interrupted time-series design

## References

ABS, 2011. Australian Statistical Geography Standard (ASGS). Australian Bureau of Statistics (ABS).https://www.abs.gov.au/websitedbs/d3310114.nsf/home/australian+statistical+geography+standard+(asgs)

ACARA, 2019. NAPLAN achievement in reading, writing, language conventions and numeracy: National report for 2019. Australian Curriculum, Assessment and Reporting Authority (ACARA). https://nap.edu.au/docs/default-source/default-document-library/2019-naplan-national-report.pdf?Status=Tempandsfvrsn=2

Adoniou, M., 2014. What should teachers know about spelling?. Literacy, 48(3), 144–154. https://doi.org/10.1111/lit.12017

Babayiğit, S., Stainthorp, R., 2011. Modeling the relationships between cognitive–linguistic skills and literacy skills: New insights from a transparent orthography. Journal of Educational Psychology, 103(1), 169. https://doi.org/10.1037/a0021671

Barry, C., and Seymour, P.H., 1988. Lexical priming and sound-to-spelling contingency effects in nonword spelling . The Quarterly Journal of Experimental Psychology Section A, 40(1), 5–40. https://doi.org/10.1080/14640748808402280

Berger, E., Carroll, M., Maybery, D., Harrison, D., 2018. Disaster impacts on students and staff from a specialist, trauma-informed Australian school. Journal of Child and Adolescent Trauma, 11, 1–10. https://doi.org/10.1007/s40653-018-0228-6

Berger, E., Gao, C.X., Jonathan, B., Campbell, T.C.H., Maybery, D., Carroll, M., 2021. The impact of a mine fire and smoke event on academic outcomes for primary and secondary school students. PsyArXiv. https://doi.org/10.31234/osf.io/unms5

Berger, E., Maybery, D., Carroll, M., 2020. Children’s perspectives on the impact of the Hazelwood mine fire and subsequent smoke event. Child and Youth Care Forum, 49, 707–724. https://doi.org/10.1007/s10566-020-09551-8

Bernal, J.L., Cummins, S., Gasparrini, A., 2017. Interrupted time series regression for the evaluation of public health interventions: A tutorial. Int J Epidemiol, 46(1), 348–355. https://doi.org/10.1093/ije/dyw098

Bosman, A. M., Van Orden, G. C., 1997. Why spelling is more difficult than reading. Learning to Spell: Research, Theory, and Practice Across Languages, 10, 173–194.

Bowman, S., McKinstry, C., McGorry, P., 2017. Youth mental ill health and secondary school completion in Australia: Time to act. Early Intervention in Psychiatry, 11(4), 277–289. https://doi.org/10.1111/eip.12357

Broder, J.C., Gao, C.X., Campbell, T.C.H., Berger, E., Maybery, D., McFarlane, A., Tsoutsoulis, J., Ikin, J., Abramson, M.J., Sim, M.R., Walker, J., Luhar, A., Carroll, M. (2020). The factors associated with distress following exposure to smoke from an extended coal mine fire. Environmental Pollution, 266(Pt 2), 115131. https://doi.org/10.1016/j.envpol.2020.115131

Buckley, T.C., Blanchard, E.B., Neill, W.T., 2000. Information processing and PTSD: A review of the empirical literature. Clinical Psychology Review, 20(8), 1041–1065.

Caffo, E., Forresi, B., Lievers, L. S., 2005. Impact, psychological sequelae and management of trauma affecting children and adolescents. Current Opinion in Psychiatry, 18(4), 422–428. https://doi.org/10.1097/01.yco.0000172062.01520.ac

Clifford, A., Lang, L., Chen, R., Anstey, K.J., Seaton, A. (2016). Exposure to air pollution and cognitive functioning across the life course–a systematic literature review. Environmental Research, 147, 383–398. https://doi.org/10.1016/j.envres.2016.01.018

CRED, 2019. CRED Crunch 58 - Disaster Year in Review. Centre for Research on the Epidemiology of Disasters (CRED). https://cred.be/sites/default/files/CC58.pdf

De Bellis, M. D., Zisk, A., 2014. The biological effects of childhood trauma. Child and Adolescent Psychiatric Clinics, 23(2), 185–222. https://doi.org/10.1016/j.chc.2014.01.002

Dimitriadis, C., Gao, C.X., Ikin, J.F., Wolfe, R., Gabbe, B.J., Sim, M.R., Abramson, M.J., Guo, Y., 2021. Exposure to mine fire related particulate matter and mortality: A time series analysis from the Hazelwood Health Study. Chemosphere. 285, 131351. https://doi.org/10.1016/j.chemosphere.2021.131351

DTF, 2015. The 2015-16 Victorian Budget For Families: Rural and Regional Budget Information Paper. Department of Treasury and Finance (DTF), Victorian Government. https://www.dtf.vic.gov.au/sites/default/files/2018-01/state-budget-rural-and-regional-bip-2015-16_0.pdf

Edwards, V.J., Holden, G.W., Felitti, V.J., Anda, R.F. (2003). Relationship between multiple forms of childhood maltreatment and adult mental health in community respondents: Results from the adverse childhood experiences study. American Journal of Psychiatry, 160(8), 1453–1460. https://doi.org/10.1176/appi.ajp.160.8.1453

Forns, J., Dadvand, P., Esnaola, M., Alvarez-Pedrerol, M., López-Vicente, M., Garcia-Esteban, R., Cirach, M., Basagaña, X., Guxens, M., Sunyer, J., 2017. Longitudinal association between air pollution exposure at school and cognitive development in school children over a period of 3.5 years. Environmental Research, 159, 416–421. https://doi.org/10.1016/j.envres.2017.08.031

Gao, C.X., Dimitriadis, C., Ikin, J., Dipnall, J.F., Wolfe, R., Sim, M.R., Smith, K., Cope, M., Abramson, M.J., Guo, Y. (2020). Impact of exposure to mine fire emitted PM_2.5_ on ambulance attendances: A time series analysis from the Hazelwood Health Study. Environmental Research, 196, 110402. https://doi.org/10.1016/j.envres.2020.110402

Gelman, A., Simpson, D., Betancourt, M., 2017. The prior can often only be understood in the context of the likelihood. Entropy, 19(10), 555. https://doi.org/10.3390/e19100555

Gibbs, L., Nursey, J., Cook, J., Ireton, G., Alkemade, N., Roberts, M., Gallagher, H.C., Bryant, R., Block, K., Molyneaux, R., Forbes, D., 2019. Delayed disaster impacts on academic performance of primary school children. Child Development, 90(4), 1402–1412. https://doi.org/10.1111/cdev.13200

Graham, S., Santangelo, T., 2014. Does spelling instruction make students better spellers, readers, and writers? A meta-analytic review. Reading and Writing, 27(9), 1703–1743.

Grievink, L., van der Velden, P. G., Yzermans, C. J., Roorda, J., Stellato, R.K., 2006. The importance of estimating selection bias on prevalence estimates shortly after a disaster. Annals of Epidemiology, 16(10), 782–788. https://doi.org/10.1016/j.annepidem.2006.04.008

Guha-Sapir, D., Hoyois, P., Wallemacq, P., Below, R., 2016. Annual disaster statistical review 2016: The numbers and trends. Centre for Research on the Epidemiology of Disasters. https://www.emdat.be/sites/default/files/adsr_2016.pdf

Hoffman, M. D., Gelman, A. (2014). The No-U-Turn sampler: adaptively setting path lengths in Hamiltonian Monte Carlo. Journal of Machine Learning Research, 15(1), 1593–1623. https://jmlr.org/papers/volume15/hoffman14a/hoffman14a.pdf

Hoffman, S. (2008). Preparing for disaster: Protecting the most vulnerable in emergencies. UC Davis Law Review, 42, 1491.

Holt, N. R., Gao, C.X., Borg, B. M., Brown, D., Broder, J.C., Ikin, J., Makar, A., McCrabb, T., Nilsen, K., Thompson, B.R., Abramson, M.J., 2021. Long-term impact of coal mine fire smoke on lung mechanics in exposed adults. Respirology. https://doi.org/10.1111/resp.14102

Huh, H. J., Kim, S.-Y., Yu, J. J., Chae, J.-H., 2014. Childhood trauma and adult interpersonal relationship problems in patients with depression and anxiety disorders. Annals of General Psychiatry, 13(1), 26. https://doi.org/10.1186/s12991-014-0026-y

IGEM. (2017). Hazelwood Mine Fire Inquiry: Implementation of recommendations and affirmations. Inspector-General for Emergency Management (IGEM), Department of Justice and Regulation, Victorian Government. https://www.vic.gov.au/sites/default/files/2019-02/Hazelwood-Mine-Fire-Inquiry-Implementation-Annual-Report-2017.pdf

Ikin, J., Carroll, M., Walker, J., Borg, B., Brown, D., Cope, M., Del Monaco, A., Dennekamp, M., Dimitriadis, C., Gao, C., Guo, Y., Johnston, F., Liew, D., Maybery, D., Thompson, B., Sim, M., M., A., 2020. Cohort Profile: The Hazelwood Health Study Adult Cohort. International Journal of Epidemiology. https://doi.org/10.1093/ije/dyaa083

Jaycox, L. H., Tanielian, T. L., Sharma, P., Morse, L., Clum, G., Stein, B. D., 2007. Schools’ mental health responses after hurricanes Katrina and Rita. Psychiatric Services, 58(10), 1339–1343. https://doi.org/10.1176/ps.2007.58.10.1339

Johnson, A.L., Dipnall, J.F., Dennekamp, M., Williamson, G.J., Gao, C.X., Carroll, M.T., Dimitriadis, C., Ikin, J.F., Johnston, F.H., McFarlane, A.C., Sim, M.R, Stub, D.A., Abramson, M.J., Guo, Y., 2019. Fine particulate matter exposure and medication dispensing during and after a coal mine fire: A time series analysis from the Hazelwood Health Study. Environmental Pollution, 246, 1027–1035. https://doi.org/10.1016/j.envpol.2018.12.085

Kar, N., 2009. Psychological impact of disasters on children: Review of assessment and interventions. World Journal of Pediatrics, 5(1), 5–11. https://doi.org/10.1007/s12519-009-0001-x

Kar, N., Bastia, B.K., 2006. Post-traumatic stress disorder, depression and generalised anxiety disorder in adolescents after a natural disaster: A study of comorbidity. Clinical Practice and Epidemiology in Mental Health, 2(1), 17. https://doi.org/10.1186/1745-0179-2-17

Kousky, C. (2016). Impacts of natural disasters on children. The Future of Children, 73–92.

Lubit, R., Rovine, D., Defrancisci, L., Eth, S., 2003. Impact of trauma on children. Journal of Psychiatric Practice, 9(2), 128–138. https://doi.org/10.1097/00131746-200303000-00004

Luhar, A.K., Emmerson, K.M., Reisen, F., Williamson, G. J., Cope, M.E., 2020. Modelling smoke distribution in the vicinity of a large and prolonged fire from an open-cut coal mine. Atmospheric Environment, 229, 117471. https://doi.org/10.1016/j.atmosenv.2020.117471

Marcotte, D.E., 2017. Something in the air? Air quality and children’s educational outcomes. Economics of Education Review, 56, 141–151. https://doi.org/10.1016/j.econedurev.2016.12.003

Margolis, A.E., Ramphal, B., Pagliaccio, D., Banker, S., Selmanovic, E., Thomas, L., Factor-Litvak, P., Perera, F., Peterson, B. S., Rundle, A., Herbstman, J., Goldsmith, J., Rauh, V., 2021. Prenatal exposure to air pollution is associated with childhood inhibitory control and adolescent academic achievement. Environmental Research, 111570. https://doi.org/10.1016/j.envres.2021.111570

Masten, A.S., Motti-Stefanidi, F., 2020. Multisystem resilience for children and youth in disaster: Reflections in the context of COVID-19. Adversity and Resilience Science, 1(2), 95–106. https://doi.org/10.1007/s42844-020-00010-w

Maybery, D., Jones, R., Dipnall, J. F., Berger, E., Campbell, T., McFarlane, A., Carroll, M., 2020. A mixed-methods study of psychological distress following an environmental catastrophe: the case of the Hazelwood open-cut coalmine fire in Australia. Anxiety, Stress, Coping, 33(2), 216–230. https://doi.org/10.1080/10615806.2019.1695523

McFarlane, A.C., Clayer, J., Bookless, C., 1997. Psychiatric morbidity following a natural disaster: An Australian bushfire. Social Psychiatry and Psychiatric Epidemiology, 32(5), 261–268. https://doi.org/10.1007/BF00789038

McFarlane, A.C., Policansky, S.K., Irwin, C., 1987. A longitudinal study of the psychological morbidity in children due to a natural disaster. Psychological Medicine, 17(3), 727–738. https://doi.org/10.1017/S0033291700025964

McLaughlin, K.A., Fairbank, J.A., Gruber, M.J., Jones, R.T., Lakoma, M.D., Pfefferbaum, B., Sampson, N.A., Kessler, R.C., 2009. Serious emotional disturbance among youths exposed to hurricane Katrina 2 years postdisaster. Journal of the American Academy of Child and Adolescent Psychiatry, 48(11), 1069–1078. https://doi.org/10.1097/CHI.0b013e3181b76697

Melody, S.M., Wheeler, A.J., Dalton, M., Williamson, G.J., Negishi, K., Willis, G., Shao, J., Zhao, B., Chappell, K., Wills, K., Reeves, M., Emmerson, K. M., Ford, J., Dennekamp, M., Foong, R. E., Abramson, M. J., Ikin, J., Walker, J., Venn, A., … Johnston, F., 2020. Cohort Profile: The Hazelwood Health Study Latrobe Early Life Follow-Up (ELF) Study. International Journal of Epidemiology, 49(6), 1779–1780. https://doi.org/10.1093/ije/dyaa136

Norris, F.H., Friedman, M.J., Watson, P.J., Byrne, C.M., Diaz, E., Kaniasty, K., 2002. 60,000 disaster victims speak: Part I. An empirical review of the empirical literature, 1981-2001. Psychiatry, 65(3), 207–239. https://doi.org/10.1521/psyc.65.3.207.20173

North, C. S., Pfefferbaum, B., 2013. Mental Health Response to Community Disasters: A Systematic Review. JAMA, 310(5), 507–518. https://doi.org/10.1001/jama.2013.107799

Peek, L., 2008. Children and disasters: Understanding vulnerability, developing capacities, and promoting resilience—an introduction. Children Youth and Environments, 18(1), 1–29. https://www.jstor.org/stable/10.7721/chilyoutenvi.18.1.0001

Peek, L., Abramson, D., Cox, R., Fothergill, A., Tobin, J., 2018. Children and disasters in: Handbook of disaster research [Book]. Springer, Cham.

Peek, L., Richardson, K., 2010. In their own words: Displaced children’s educational recovery needs after Hurricane Katrina. Disaster Med Public Health Prep, 4 Suppl 1, S63–70. https://doi.org/10.1001/dmp.2010.10060910

Perez-Pereira, M., Tinajero, C., Rodriguez, M.S., Peralbo, M., Sabucedo, J.M., 2012. Academic effects of the prestige oil spill disaster. Span J Psychol, 15(3), 1055–1068.

Perfect, M.M., Turley, M.R., Carlson, J.S., Yohanna, J., Saint Gilles, M.P., 2016. School-related outcomes of traumatic event exposure and traumatic stress symptoms in students: A systematic review of research from 1990 to 2015. School Mental Health: A Multidisciplinary Research and Practice Journal, 8(1), 7–43. https://doi.org/10.1007/s12310-016-9175-2

Perry, B. D., 2006) Applying principles of neurodevelopment to clinical work with maltreated and traumatized children: The neurosequential model of therapeutics [Book Section]. In Working with traumatized youth in child welfare. (pp. 27–52). The Guilford Press.

Pietro, G. D., 2018. The academic impact of natural disasters: Evidence from l’Aquila earthquake. Education Economics, 26(1), 62–77. https://doi.org/10.1080/09645292.2017.1394984

Pullins, L.G., McCammon, S.L., Lamson, A.S., Wuensch, K.L., Mega, L., 2005. School-based post-flood screening and evaluation: Findings and challenges in one community. Stress, Trauma, and Crisis, 8(4), 229–249. https://doi.org/10.1080/15434610500406343

Reisen, F., Gillett, R., Choi, J., Fisher, G., Torre, P., 2017. Characteristics of an open-cut coal mine fire pollution event. Atmospheric Environment, 151, 140–151. https://doi.org/10.1016/j.atmosenv.2016.12.015

Romano, E., Babchishin, L., Marquis, R., Fréchette, S., 2015. Childhood maltreatment and educational outcomes. Trauma, Violence, Abuse, 16(4), 418–437. https://doi.org/10.1177/1524838014537908

Salloum, A., Overstreet, S., 2012. Grief and trauma intervention for children after disaster: Exploring coping skills versus trauma narration. Behaviour Research and Therapy, 50(3), 169–179. https://doi.org/10.1016/j.brat.2012.01.001

Samuelson, K.W., Krueger, C.E., Burnett, C., Wilson, C.K., 2010. Neuropsychological functioning in children with posttraumatic stress disorder. Child Neuropsychology, 16(2), 119–133. https://doi.org/10.1080/09297040903190782

Saylor, C. F. (1993). Children and disasters [Book]. New York: Plenum Press.

Shao, J., Zosky, G.R., Hall, G.L., Wheeler, A. ., Dharmage, S., Melody, S., Dalton, M., Foong, R. E., O’Sullivan, T., Williamson, G.J., Chappell, K., Abramson, M.J., Johnston, F.H., 2020. Early life exposure to coal mine fire smoke emissions and altered lung function in young children. Respirology, 25(2), 198–205. https://doi.org/10.1111/resp.13617

Silverman, W.K., La Greca, A.M., 2002. Children experiencing disasters: Definitions, reactions, and predictors of outcomes. In Helping children cope with disasters and terrorism. (pp. 11–33). American Psychological Association. https://doi.org/10.1037/10454-001

Siriwardhana, C., Pannala, G., Siribaddana, S., Sumathipala, A., Stewart, R., 2013. Impact of exposure to conflict, tsunami and mental disorders on school absenteeism: Findings from a national sample of Sri Lankan children aged 12-17 years. BMC Public Health, 13, 560. https://doi.org/10.1186/1471-2458-13-560

Stan Development Team, 2020. RStan: The R interface to Stan. https://cran.r-project.org/web/packages/rstan/rstan.pdf

Stan Development Team, 2020. Stan user’s guide, version 2.25. https://mc-stan.org/

Tang, W., Zhao, J., Lu, Y., Yan, T., Wang, L., Zhang, J., Xu, J., 2017. Mental health problems among children and adolescents experiencing two major earthquakes in remote mountainous regions: A longitudinal study. Comprehensive Psychiatry, 72, 66–73. https://doi.org/10.1016/j.comppsych.2016.09.004

Terasaka, A., Tachibana, Y., Okuyama, M., Igarashi, T., 2015. Post-traumatic stress disorder in children following natural disasters: A systematic review of the long-term follow-up studies. International Journal of Child, Youth and Family Studies, 6(1), 111–133. https://doi.org/10.18357/ijcyfs.61201513481

Treiman, R., 2018. Teaching and learning spelling. Child Development Perspectives, 12(4), 235–239. https://doi.org/10.1111/cdep.12292

UNDRR, 2020. The human cost of disasters: An overview of the last 20 years (2000-2019). UN Office for Disaster Risk Reduction (UNDRR). https://www.undrr.org/sites/default/files/inline-files/Human%20Cost%20of%20Disasters%202000-2019%20FINAL.pdf

van Houwelingen, H.C., Arends, L.R. and Stijnen, T., 2002. Advanced methods in meta-analysis: multivariate approach and meta-regression. Statist. Med., 21, 589–624. https://doi.org/10.1002/sim.1040

VCOSS, 2015. Submission to the 2015 Hazelwood Mine Fire Inquiry [Report]. Victorian Council of Social Services (VCOSS). http://hazelwoodinquiry.vic.gov.au/wp-content/uploads/2015/08/Victorian-Council-of-Social-Service-PDF-483KB.pdf

Xu, R., Yu, P., Abramson, M. J., Johnston, F. H., Samet, J.M., Bell, M.L., Haines, A., Ebi, K. L., Li, S., Guo, Y., 2020. Wildfires, global climate change, and human health. New England Journal of Medicine, 383(22), 2173–2181. https://doi.org/10.1056/NEJMsr2028985

Yell, S., Duffy, M., Whyte, S., Walker, L., Carroll, M., Walker, J., 2019a. Hazelwood Health Study Community Wellbeing Stream Report Volume 1: Community perceptions of the impact of the smoke event on community wellbeing and of the effectiveness of communication during and after the smoke event. https://hazelwoodhealthstudy.org.au/data/assets/pdf_file/0018/2052540/CW-Report-Volume-1_v2.0.pdf

Yell, S., Duffy, M., Whyte, S., Walker, L., Carroll, M., Walker, J., 2019b. Hazelwood Health Study Community Wellbeing Stream Report Volume 2: Community perceptions of the effectiveness of community rebuilding activities. https://hazelwoodhealthstudy.org.au/data/assets/pdf_file/0009/2059236/CW-Report-Volume-2_version-1.0.pdf

Yelland, C., Robinson, P., Lock, C., La Greca, A. M., Kokegei, B., Ridgway, V., Lai, B., 2010. Bushfire impact on youth. Journal of Traumatic Stress, 23(2), 274–277. https://doi.org/10.1002/jts.20521

